# Scalable Causal-Interpretable Machine Learning for Cancer Prescreening Using Electronic Health Records

**DOI:** 10.64898/2026.09.21.26363536

**Authors:** Shuaijie Zhang, Fuzhong Xue

**Affiliations:** Beijing Key Laboratory of Topological Statistics and Applications for Complex Systems, Beijing Institute of Mathematical Sciences and Applications, Beijing 101408, China; Yau Mathematical Sciences Center, Tsinghua University, Beijing 100084, China; Department of Medical Dataology, School of Public Health, Cheeloo College of Medicine, Shandong University, Jinan 250012, China; National Institute of Health and Medical Big Data, Jinan 250012, China; Qilu Hospital, Cheeloo College of Medicine, Shandong University, Jinan 250012, China

**Keywords:** Causal machine learning, Cancer prescreening, Large-scale data, Medicine decision support

## Abstract

Electronic health records provide a source of real-world information for disease risk prediction, but many machine learning models learn implicit associations that are difficult to inspect or use for clinical reasoning. Causal Bayesian networks (CBNs) offer a form of causal-interpretable machine learning by representing conditional dependencies, putative directional relations, and probabilistic evidence propagation in a directed acyclic graph. However, conventional CBN structure learning becomes unstable and computationally expensive when applied to large-scale EHR data. To address this limitation, we propose UPEBNL, a scalable framework for causal-interpretable cancer prescreening based on parallel CBN learning. UPEBNL integrates adaptive data slicing, quality-aware structure aggregation, and global DAG construction to learn stable and interpretable dependency structures from large-scale observational data. We evaluated UPEBNL in high-dimensional and multi-million-sample simulations and applied it to EHR-based prescreening for esophageal and colorectal cancer. In simulations, UPEBNL improved structural recovery accuracy by nearly 40% and achieved up to a 221.28-fold speedup over conventional CBN learning strategies. For cancer risk prediction, the learned CBNs provided interpretable evidence paths and achieved validation AUCs of 0.8171 for esophageal cancer and 0.784 for colorectal cancer. Calibration and decision curve analyses further supported the reliability and clinical utility of the models. These findings suggest that scalable CBN learning can support interpretable cancer prescreening from large-scale EHR data.

## 1. Introduction

Large language models and deep learning models have shown strong representational capacity in large-scale data modeling and have been used in real-world medicine decision settings [1, 2]. However, the knowledge learned by these models is often stored implicitly in parameters or embeddings, without an explicit structure that supports reasoning [2, 3, 4]. As a result, these models often provide limited support for intervention analysis and trustworthy decision support [3, 5]. Importantly, large-scale real-world observational data contain rich latent causal relations [6, 5, 7]. Discovering these relations from large-scale observational data and converting them into structured causal knowledge is an important step toward interpretable and trustworthy decision support [7, 8].

Causal Bayesian networks (CBNs) provide a suitable tool for structured causal knowledge representation [8, 9]. Through a directed acyclic graph, a CBN explicitly encodes causal directions and transmission paths hidden in data, allowing the learned knowledge to be queried and used for probabilistic reasoning [8, 9]. This property makes CBNs suitable for knowledge-driven decision support. For example, in medical risk prediction, such structured representations can help identify evidence paths related to disease outcomes and improve the interpretability of prediction results [10].

However, real-world data often involve large sample sizes, high dimensionality, and complex dependencies [6, 7]. As the number of variables and data complexity increase, potential causal paths and candidate edges grow rapidly [8, 11]. Traditional CBN learning algorithms therefore struggle to identify causal relations from large variable combinations in a stable and accurate manner [8, 12, 13]. This makes it difficult to organize these relations as structured causal knowledge, limiting the use of existing CBN methods in large-scale real-world decision settings.

To address this problem, this paper proposes UPEBNL, a causal knowledge discovery framework based on parallel CBN learning, for learning structured causal knowledge from large-scale observational data. UPEBNL uses adaptive data slicing, quality-aware structure aggregation, and global DAG construction to learn causal knowledge structures that can be used for probabilistic reasoning and decision support in complex data scenarios.

The main contributions are as follows.

1. UPEBNL is proposed to discover structured causal knowledge from large-scale observational data and generate a CBN representation that supports reasoning.
2. Adaptive data slicing, quality-aware structure aggregation, and global DAG construction are designed to improve the accuracy and learning efficiency of causal knowledge discovery in large-scale data.
3. UPEBNL is evaluated in high-dimensional simulations with multi-million-sample data, followed by a large-scale EHR-based cancer prediction application. The results show that UPEBNL improves causal knowledge discovery accuracy and learning efficiency, and that the learned structured causal knowledge can support interpretable cancer risk prediction.

The remainder of this paper is organized as follows. Section 2 provides an overview of related work. Section 3 presents the proposed UPEBNL framework for structured causal knowledge construction. Section 4 reports the simulation design and results. Section 5 presents the application of UPEBNL to large-scale EHR-based cancer prediction and analyzes how the learned structured causal knowledge supports interpretable risk prediction. Section 6 discusses the main findings and concludes the work.

## 2. Related work

Causal Bayesian networks (CBNs) provide a graphical representation for causal knowledge [8, 9, 14]. In a CBN, a directed acyclic graph encodes causal dependencies among variables, while conditional probability distributions specify how probabilities propagate across different variable states [9, 15]. Compared with black-box machine learning models [3, 16], CBNs can, under causal assumptions [9, 17], represent causal knowledge learned from data as causally interpretable graphs. This graph supports posterior updating for a target event given observed evidence and allows researchers to examine how effects are transmitted along network paths. In this sense, CBNs provide a graphical framework that connects causal knowledge representation, probabilistic reasoning, and decision analysis.

The structured representation of causal knowledge depends on Bayesian network structure learning. Existing structure learning methods are commonly grouped into constraint-based, score-based, and hybrid approaches [8, 18]. Constraint-based methods rely on conditional independence tests to recover the network skeleton from independence and dependence relations among variables [11, 18]. Edge directions are then inferred using local structures, such as collider patterns, and orientation rules. Representative algorithms include PC [11] and Fast IAMB [19]. By contrast, score-based methods formulate structure learning as a combinatorial optimization problem. They evaluate candidate network structures using scoring criteria such as BIC and search for high-scoring structures through strategies such as greedy search or hill climbing [20, 21]. Hybrid methods combine these two ideas by first using local dependence information or Markov blanket discovery to restrict the candidate parent sets, and then applying score-based search within the reduced space [22]. MMHC is a typical example of this class [22]. Among these methods, constraint-based approaches are more directly aligned with Pearl’s causal theory, because their learning principle is based on the correspondence between graphical separation and probabilistic independence [9, 17]. Accordingly, this paper focuses on constraint-based Bayesian network structure learning as a basis for CBN-based causal knowledge discovery.

However, applying CBN structure learning to large-scale observational data leads to a rapid increase in the number of conditional independence tests as the number of variables grows [8, 12, 23]. The complexity of conditioning sets also grows, which substantially reduces learning efficiency. Moreover, in high-dimensional settings, local testing errors may propagate through the process of causal structure identification [24]. Such errors can lead to missing edges, redundant dependencies, or incorrect orientations, thereby reducing the accuracy of the learned structure. As a result, the credibility of the resulting CBN as a causal knowledge base for reasoning and decision support may be weakened.

To address these challenges, existing studies have explored several strategies for improving CBN structure learning in large-scale settings [25, 26, 27]. One direction seeks to reduce the candidate search space by using Markov blankets to remove variables weakly related to the target node [18, 22, 28]. These methods can reduce the size of the candidate parent set for each node. Their structural quality, however, depends heavily on the stability of local conditional independence tests. If the local candidate set contains missing or redundant edges, the completeness of causal paths and the reliability of edge orientations in the global CBN may be affected. A second direction focuses on computational scalability in causal structure learning. Parallel methods usually distribute conditional independence tests or candidate edge screening across multi-core or distributed computing environments to improve learning efficiency on large-scale data. For example, Madsen et al. [29], Le et al. [23], and Yang et al. [30] developed parallel structure learning approaches, allowing parts of conditional independence testing or edge screening to be executed in parallel. These methods improve the feasibility of causal structure learning in high-dimensional settings. Nevertheless, their applicability to large-scale observational data remains limited by sample-size constraints and substantial computational requirements, especially when the data contain millions of observations.

Accordingly, large-scale CBN-based causal knowledge learning requires both structural accuracy and computational efficiency, allowing the resulting network to support posterior probability updating and decision-oriented analysis. These requirements motivate the development of the UPEBNL framework proposed in this paper.

## 3. Proposed methodology

The proposed UPEBNL framework provides a scalable approach for causal knowledge discovery from large-scale observational data by integrating adaptive data slicing, quality-aware structure aggregation, and global DAG construction. UPEBNL learns local CBN structures from appropriately sized data slices and integrates them into a global causal knowledge structure. Adaptive slicing determines data subsets that remain suitable for local structure learning while reducing the burden of large-scale conditional independence testing. Quality-aware aggregation assigns different contributions to locally learned structures according to their structural quality, thereby reducing the influence of unstable local results. Global DAG construction resolves directional conflicts and produces a coherent CBN that can be used for probabilistic reasoning and decision support. The framework of the proposed methodology is presented in Fig. 1 and Fig. 2.

**Fig. 1:**
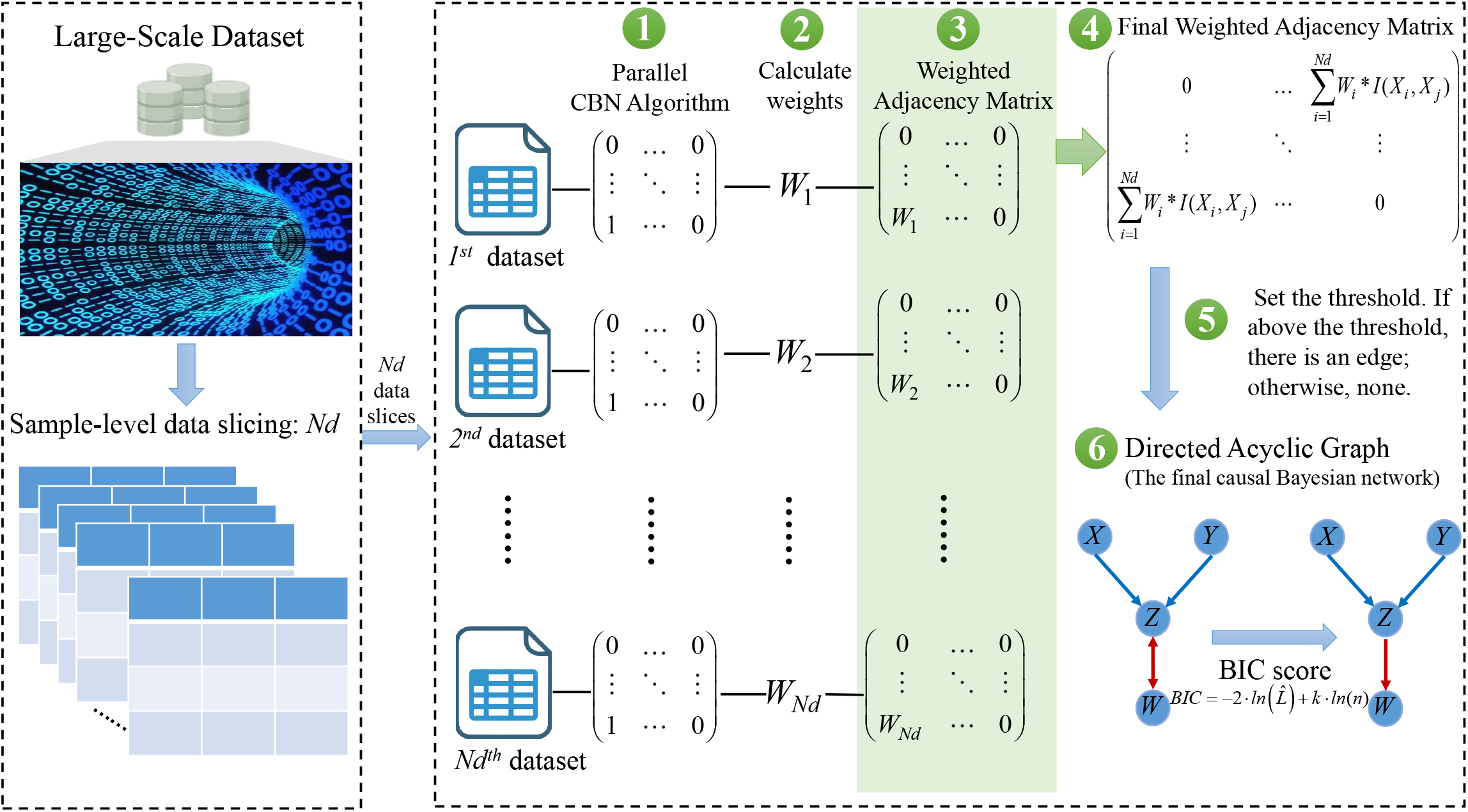
Flowchart of the UPEBNL algorithm for learning structured causal knowledge.

**Fig. 2:**
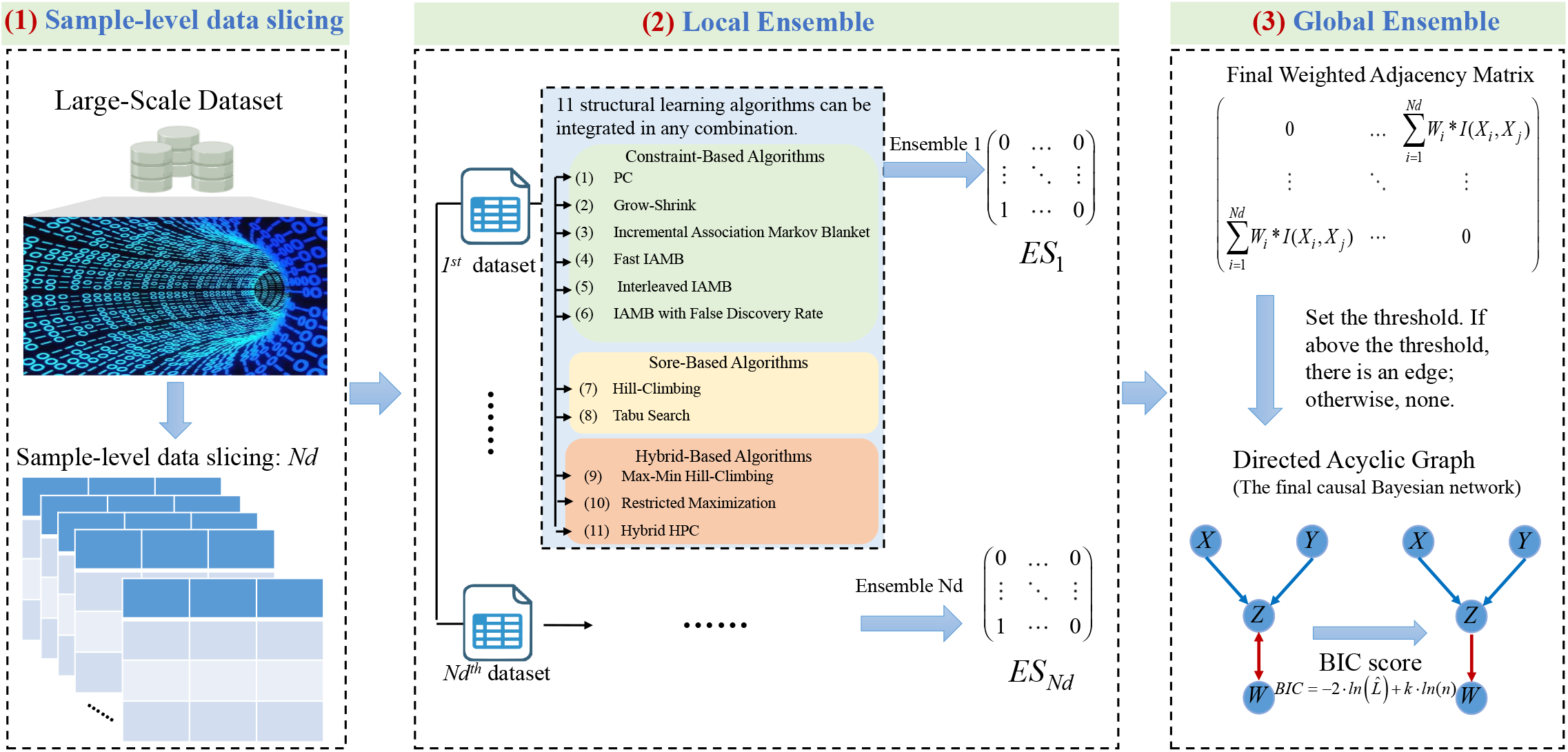
Flowchart of the extended UPEBNL algorithm for integrating multiple knowledge structure learners.

### 3.1. Preliminaries

#### Definition 1

***Average markov blanket size*** [25]. In a CBN *B* with *p* nodes, the Average Markov Blanket Size (AMBS) is a measure that represents the average number of nodes in the MB of each node within the network. Formally, the Markov Blanket Size (MBS) of a node *X*_*i*_ is the number of nodes in its MB. The AMBS is calculated by summing the MBS of all nodes in the network and then dividing by the total number of nodes *p*.

Mathematically, the AMBS is defined as follows:

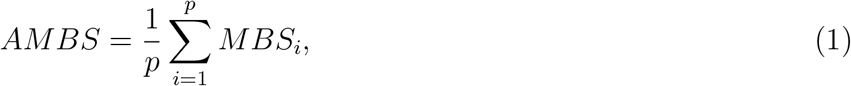

where *MBS*_*i*_ represents the MBS of node *X*_*i*_, and *p* is the total number of nodes in the CBN.

#### Definition 2

***Bayesian Dirichlet equivalent uniform score*** [31, 32] *Bayesian Dirichlet equivalent uniform (BDeu) score is used to evaluate the goodness-of-fit of a CBN given a specific dataset and structure. It combines prior information with the likelihood of the data. According to the Markov properties of CBNs, two Markov equivalent CBNs have the same BDeu score*.

*The BDeu formula is as follows:*

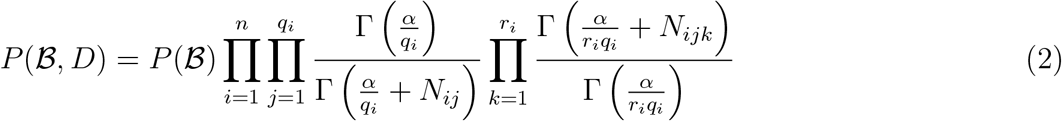

where *B* denotes the structure of the CBN; *D* represents the dataset; *n* is the number of variables in the network (assuming the variables in *D* are all the variables in the network); *q*_*i*_ is the number of parent configurations for variable *X*_*i*_; *r*_*i*_ is the number of possible values that variable *X*_*i*_ can take;*α* is a hyperparameter reflecting the prior equivalent sample size; *N*_*ij*_ is the number of instances in the dataset where the parents of node *X*_*i*_ are in the *j*-th configuration; *N*_*ijk*_ is the number of instances where node *X*_*i*_ is in the *k*-th state and its parents are in the *j*-th configuration; Γ is the gamma function.

#### Definition 3

***Edge strength*** [25]. Edge Strength (ES) is a metric used to evaluate the average contribution of each edge in a CBN. It measures the quality of the CBN by combining the score of the network and the network complexity, which is determined by the number of edges and the amount of data. Given a dataset *D* containing *N* samples and a CBN *B* containing *M* edges, the ES is calculated using the following formula:

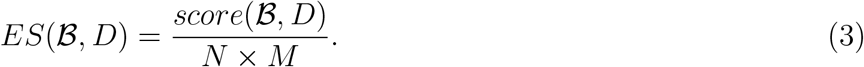

In this study, the BDeu score is utilized to calculate the ES.

#### Definition 4

***d-separation*** [13]. Let *X, Y* , and *Z* be disjoint sets of nodes in a DAG, and let *π* be a path. **W** is a subset of *V*. The path *π* is *blocked* [33] by **W** if and only if *π* contains: (1) a fork *X* ← *Y* → *Z* or a chain *X* → *Y* → *Z* such that the middle vertex *Y* is in **W**, or (2) a collider *X* → *Y* ← *Z* such that the middle vertex *Y* , or any descendant of it, is not in **W**. The set **W** d-separates *X* from *Z* if it blocks every path between *X* and *Z*.

CBN structure learning typically uses CI tests to identify dependencies and independencies among variables. Given a complete dataset *D* with all discrete variables, CI tests are commonly performed using the *G*^2^ statistic [17]. The formula for the *G*^2^ statistic is as follows [34]:

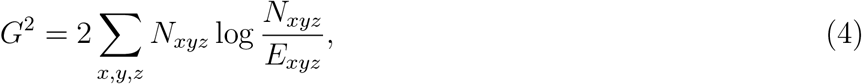

where *N*_*xyz*_ is the number of samples in the dataset which satisfies *V*_*i*_ = *x, V*_*j*_ = *y*, and *V*_*k*_ = *z*. The expected frequency *E*_*xyz*_ is defined as

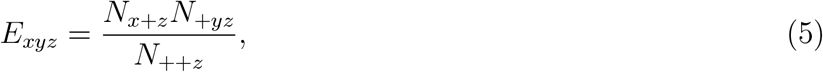

where *N*_*x*+*z*_, *N*_+*yz*_, and *N*_++*z*_ are the marginal frequencies,*N*_*x*+*z*_ = ∑ _*y*_ *N*_*xyz*_, *N*_+*yz*_ = ∑ _*x*_ *N*_*xyz*_, and *N*_++*z*_ = ∑_*xy*_ *N* _*xyz*_.

The *G*^2^ statistic asymptotically follows a *χ*^2^ distribution with degrees of freedom (|*V*_*i*_| − 1)(|*V*_*j*_| − 1), where | · | denotes the number of possible values of the variable. The p-value of the *χ*^2^ distribution can be calculated based on the *G*^2^ statistic. The conclusion of independence is determined by comparing the p-value with the significance level *α*. If the p-value is lower than the significance threshold *α*, the null hypothesis is rejected, and ¬*CI*(*V*_*i*_, *V*_*j*_ | *{V*_*k*_*}*) is determined to be true. If the p-value is greater than *α*, the independence hypothesis *CI*(*V*_*i*_, *V*_*j*_ | *{V*_*k*_*}*) is accepted.

### 3.2. UPEBNL for causal knowledge discovery

UPEBNL learns causal knowledge in a divide-and-integrate manner. The original dataset is divided into data slices suitable for local structure learning, and each slice is processed in parallel to obtain a candidate CBN. These candidate structures are then integrated through weighted ensemble learning to form a global causal knowledge structure. The procedure of UPEBNL for learning a single CBN is illustrated in Fig. 1.

#### Stage I: Causal-knowledge-preserving data segmentation

In UPEBNL, data segmentation divides the samples into subsets while retaining the full variable set in each subset. The slice size affects both the computational burden and the structural reliability of the causal relations learned from each subset. Large slices retain more samples but may remain costly for conditional independence testing, whereas overly small slices may weaken faithfulness and produce unstable local causal representations. Tang et al. [25] introduced the Appropriate Learning Size (ALS) for large datasets that are independently and identically distributed and conform to a DAG. ALS is defined as the minimum slice size required to preserve the DAG-related properties of the original data.

Tang et al. [25] demonstrated the existence of ALS using AMBS and ES. Let a large dataset *D*_2_ and a smaller dataset *D*_1_ be drawn from the same distribution *P* , and assume that both faithfully represent the same underlying DAG. Under this condition, 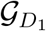 and 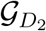 encode the same conditional independencies implied by *P*. Their learned graph structures therefore belong to the same equivalence class and may differ only in edge orientation. According to the definitions of AMBS (Eq. (1)) and ES (Eq. (3)), both metrics are invariant to edge direction. Therefore, a threshold exists at which the differences in AMBS and ES between the two graphs converge.

In this paper, we use *ALS_with_PK*, an extension of the ALS method proposed by Tang et al. [25]. When prior causal constraints are available, *ALS_with_PK* incorporates them into the segmentation process so that each data slice is more likely to retain the causal relations needed for subsequent knowledge discovery.

#### Stage II: Quality-aware causal structure aggregation

CBNs learned from different data slices may contribute unequally to causal knowledge construction. UPEBNL therefore evaluates the structure learned from each slice and integrates these structures using quality-aware weights. The CBN causal structure learning algorithm is applied to the *N*_*d*_ data slices in parallel, with one candidate CBN learned from each slice. To quantify the contribution of the CBN learned from the *k*-th data slice *D*_*k*_, we calculate its weight using ES as

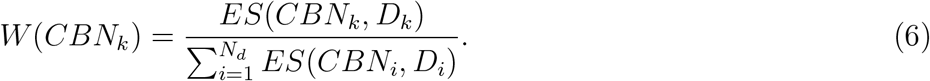

A larger value of *W* (*CBN*_*k*_) indicates a greater contribution of the corresponding CBN to the aggregated causal structure. Let 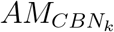 denote the adjacency matrix of *CBN*_*k*_, where an entry of 1 indicates the presence of an edge and an entry of 0 indicates its absence. The weighted adjacency matrix of *CBN*_*k*_ is then obtained by multiplying each entry of 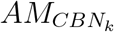 by its corresponding weight:

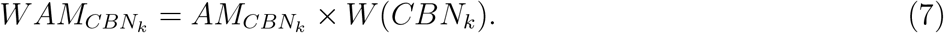

The final weighted adjacency matrix is constructed by summing the weighted adjacency matrices over all *N*_*d*_ candidate CBNs:

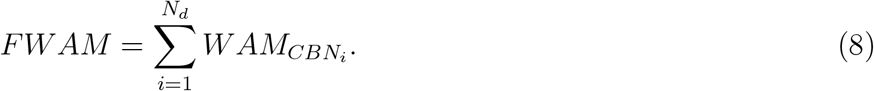

To obtain the final adjacency matrix, a threshold is defined as *γ* = *T ×* min(*W* (*CBN*_*k*_)), where *T* specifies the minimum required level of support across the weighted networks. Each element of *FWAM* is then compared with *γ*. Elements greater than or equal to *γ* are set to 1, indicating the presence of an edge, whereas elements below *γ* are set to 0. This thresholding step yields the adjacency matrix of the integrated network. At this stage, the integrated network may still contain bidirectional edges.

#### Stage III: Global DAG restoration for probabilistic reasoning

Prediction with CBNs requires the network to be a DAG. If bidirectional edges remain from the previous stage, their orientations are further determined according to the Bayesian information criterion (BIC) [20, 35], yielding the final DAG. A lower BIC value indicates a better-fitting model. BIC is formally defined as

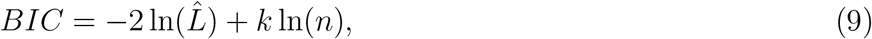

where 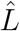 denotes the maximized likelihood of the CBN, *n* is the sample size, and *k* is the number of parameters estimated in the CBN.

Algorithm 1 gives the pseudo-code of the UPEBNL algorithm for causal knowledge discovery.

##### Algorithm 1

The UPEBNL Algorithm for causal knowledge discovery

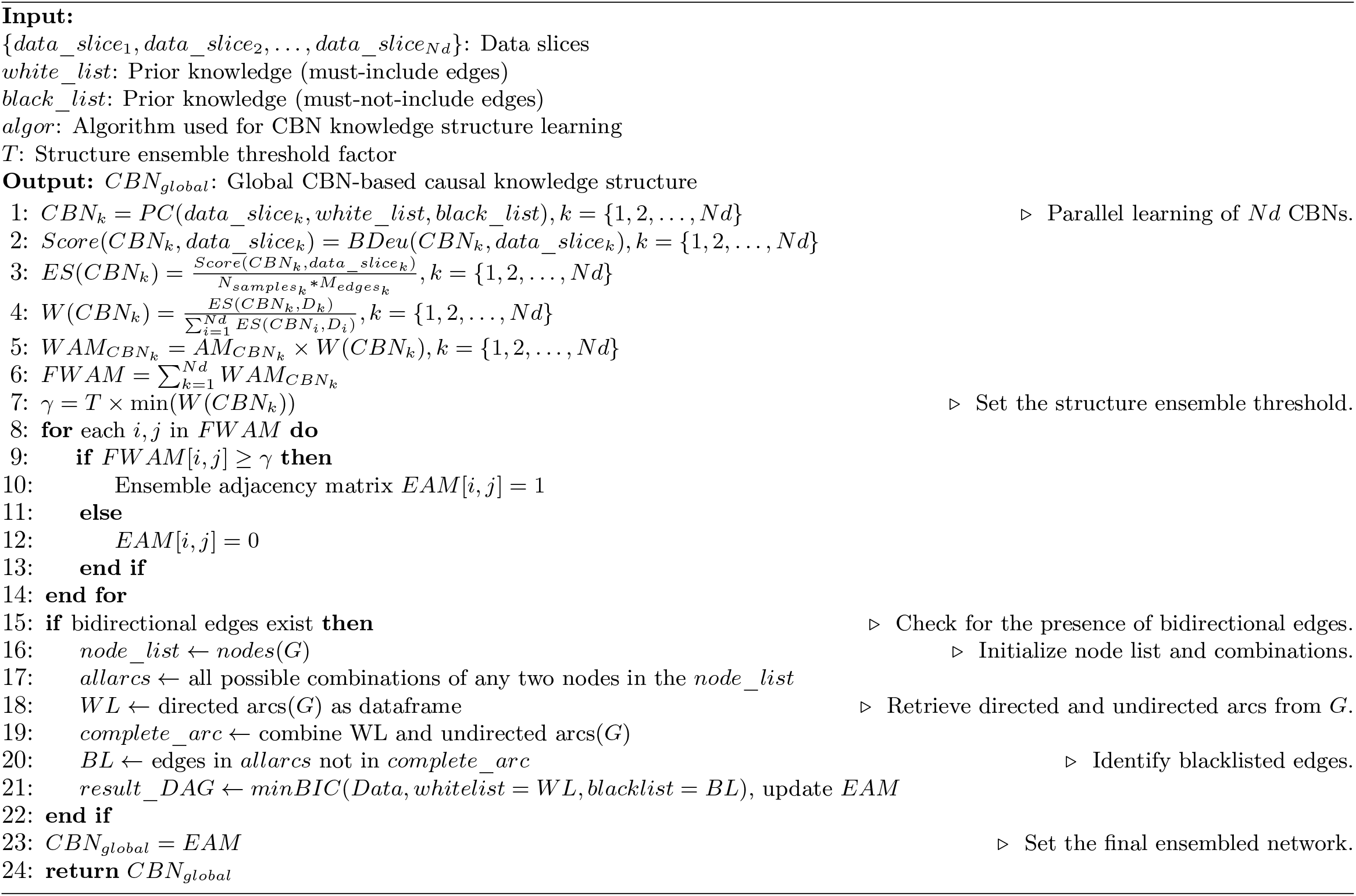

### 3.3. Generalization to multiple knowledge structure learners

We extend UPEBNL to multiple knowledge structure learning algorithms, including constraint-based, score-based, and hybrid methods, to form consistent causal knowledge from multi-algorithm results. In this parallel multi-algorithm setting, UPEBNL follows three steps: dataset slicing, local ensemble, and global ensemble, as shown in Fig. 2.

For data slicing, the slice size is no longer determined from the result of a single learner. During the iterative process of *ALS_with_PK*, UPEBNL first locally integrates the candidate networks obtained by different algorithms on the same data slice, and then evaluates whether the integrated result retains sufficient structured knowledge information. The pseudocode is provided in Supplementary Algorithm S1.

Local ensemble integrates the results produced by multiple algorithms on the same data slice. Different algorithms may identify different dependencies from the same data slice. The local ensemble summarizes these candidate relations into the knowledge structure corresponding to that slice, thereby reducing the influence of single-algorithm bias on the result. The pseudocode is provided in Supplementary Algorithm S2.

Global ensemble further integrates the knowledge structures obtained from different data slices. UPEBNL combines the results from all slices according to the weighted ensemble principle defined above and generates the final global CBN. This step aims to aggregate structured knowledge information distributed across different data slices into a stable and reliable overall representation. The pseudocode is provided in Supplementary Algorithm S3.

## 4. Experiments and results

### 4.1. Simulation design

To evaluate UPEBNL for structured causal knowledge discovery from large-scale data, we designed a series of simulation experiments considering sample size, number of variables, and network density. Network density reflects the complexity of potential causal dependencies. UPEBNL was evaluated with constraint-based, score-based, and hybrid structure learning algorithms to examine its applicability across different knowledge structure learning strategies.

The simulations used binary data, with all variables taking values of either 0 or 1. Different network densities were generated by adjusting the `prob` parameter in the `random.graph` function in R. A larger `prob` value produces more edges and therefore more potential causal paths to be identified. For a fixed number of variables, a denser network contains more candidate causal paths and dependency directions. Table 1 reports the number of edges under different node numbers and `prob` settings.

**Table 1:** Statistics of causal-dependency complexity under different numbers of variables.

| Number of Nodes (Variables) | Prob Parameter Value | Number of Edges |
| --- | --- | --- |
| 50 | NULL | 52 |
| 100 | NULL | 93 |
| 200 | NULL | 206 |
| 300 | NULL | 312 |
| 400 | NULL | 396 |
| 500 | NULL | 479 |
| 100 | 0.05 | 246 |
| 100 | 0.10 | 487 |
| 100 | 0.15 | 732 |
| 100 | 0.20 | 977 |
| 50 | 0.05 | 62 |
| 50 | 0.10 | 122 |
| 50 | 0.15 | 198 |
| 50 | 0.20 | 265 |
| 200 | 0.05 | 992 |
| 200 | 0.10 | 1,986 |
| 200 | 0.15 | 2,930 |
| 200 | 0.20 | 3,931 |

Notes: The parameter `prob` determines network density, which reflects the complexity of potential causal dependencies. When `prob` is set to `NULL`, it indicates a sparse dependency structure. Larger `prob` values generate more edges and therefore denser causal-dependency structures.

We first considered constraint-based algorithms. We selected the PC algorithm and the Fast IAMB algorithm for comparison. Each setting was repeated 100 times. The simulation scenarios were as follows:

- **Scenario 1:** With the sample size fixed at 1,000,000 and network density fixed at `prob=NULL`, the number of variables was set to {50, 100, 200, 300, 400, 500}. This scenario assessed the stability of causal knowledge discovery as dimensionality increased.
- **Scenario 2:** With the number of variables fixed at 100 and network density fixed at `prob=NULL`, the sample size was set to {500,000, 1,000,000, 2,000,000, 3,000,000, 4,000,000, 5,000,000}. This scenario assessed performance under multi-million-sample conditions.
- **Scenario 3:** Using the PC algorithm, we varied the number of variables, sample size, and network density to assess UPEBNL under different levels of network density.

To examine the extensibility of UPEBNL across knowledge structure learning algorithms, we also considered score-based and hybrid methods. Based on the learning performance reported in previous work [36], we selected Tabu Search [37], Restricted Maximization, and Hybrid HPC as representative algorithms. We also included a hybrid learning strategy combining the constraint-based PC algorithm with the score-based Tabu algorithm. The simulation scenario was as follows:

- **Scenario 4:** With the network density parameter set to `prob=NULL`, the number of variables was set to {50, 100, 200, 300, 400, 500}. This scenario assessed UPEBNL with score-based and hybrid algorithms.

As the reference for assessing the learned causal knowledge structures, we constructed a gold standard CBN under each simulation setting using the `random.graph` function in R. The networks learned by UPEBNL and the comparison methods were then compared with the gold standard CBN to assess the recovery of the underlying structured causal relations.

#### 4.1.1. Compared algorithms

To assess the relative performance of UPEBNL in structured causal knowledge discovery, we compared it with the baseline implementations provided by the `bnlearn` package in R and with traditional non-parallel implementations. The parallel implementations in `bnlearn` support constraint-based and score-based algorithms, but not hybrid algorithms. Therefore, for hybrid algorithms, UPEBNL was compared only with traditional non-parallel implementations. For the parallel algorithms, the experiments used 31 CPU cores. A cluster object with 31 worker nodes was created using the `makeCluster` function from the `parallel` package in R.

#### 4.1.2. Evaluation metrics

To evaluate the accuracy and learning efficiency of the learned causal knowledge structures, we used six metrics: Precision, Recall, F1 Score, False Discovery Rate (FDR), Structural Hamming Distance (SHD), and Time (mins).

- **(1) Precision:** Precision represents the proportion of causal dependencies in the learned causal knowledge structure that are correctly identified and also present in the gold standard network. The formula is:

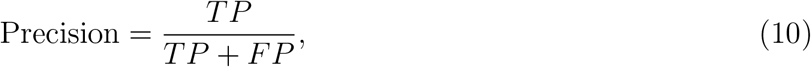

where *TP* denotes causal dependencies present in both the learned network and the gold standard network, and *FP* denotes causal dependencies present only in the learned network.
- **(2) Recall:** Recall represents the proportion of causal dependencies in the gold standard network that are correctly identified by the learning method. The formula is:

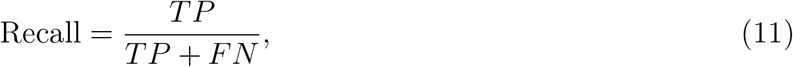

where *FN* denotes causal dependencies present only in the gold standard network but not identified by the learned network.
- **(3) F1 Score:** The F1 Score is the harmonic mean of Precision and Recall and provides an overall measure of the accuracy and completeness of causal relation identification. The formula is:

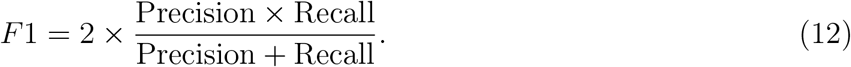
- **(4) False Discovery Rate (FDR):** FDR represents the proportion of incorrectly identified causal dependencies in the learned causal knowledge structure. The formula is:

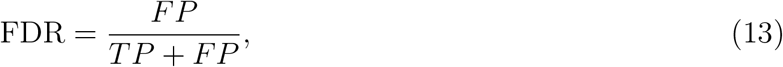

where *FP* denotes causal dependencies present in the learned network but absent from the gold standard network.
- **(5) Structural Hamming Distance (SHD):** SHD measures the minimum number of dependency insertions, deletions, and direction reversals required to transform the learned network into the gold standard network. The formula is:

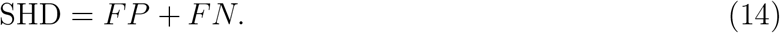

This metric reflects the structural difference between the learned causal knowledge structure and the true network. A smaller SHD indicates that the learned network is closer to the gold standard network.
- **(6) Computation Time:**

Computation Time denotes the time required to complete causal knowledge structure learning, measured in minutes.

The following subsections report the results for the different simulation scenarios. In some scenarios, the `bnlearn` parallel and non-parallel algorithms required excessive running time and did not complete all 100 simulations. These incomplete results were therefore excluded from the analysis.

### 4.2. Performance under increasing dimensionality

In the simulations with increasing dimensionality, the sample size was fixed at 1,000,000 and the network density parameter was set to `prob=NULL`. This setting evaluated whether UPEBNL could stably discover structured causal knowledge from large-scale data as the number of candidate variables increased. As shown in Fig. 3, when the number of variables increased from 50 to 500, the Precision, Recall, and F1 Score of the bnlearn parallel and traditional non-parallel algorithms decreased. Taking the PC algorithm as an example, when the number of variables was 50, the Precision of both the bnlearn parallel and non-parallel PC algorithms was approximately 0.62. When the number of variables increased to 500, Precision decreased to approximately 0.5. In contrast, UPEBNL maintained Precision, Recall, and F1 Score above 0.9 in most scenarios, indicating that it identified true causal dependencies more stably in high-dimensional settings.

**Fig. 3:**
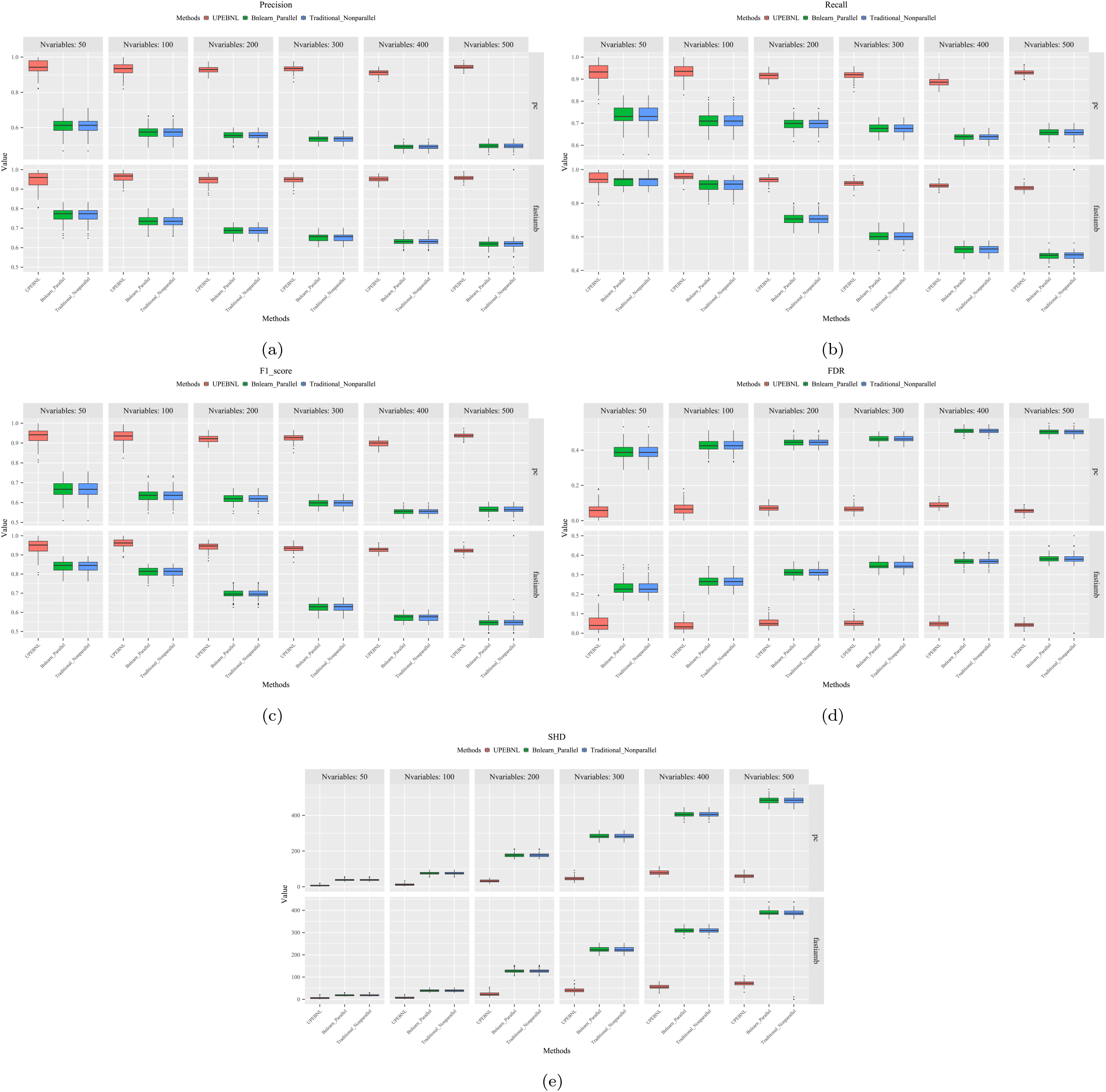
Evaluation of structured causal knowledge discovery with 1 million samples and varying numbers of variables. metrics include (a) Precision, (b) Recall, (c) F1 Score, (d) FDR, and (e) SHD. Higher Precision, Recall, and F1 Score indicate e accurate and complete identification of causal dependencies, while lower FDR and SHD indicate fewer false dependencies and ller structural deviations. `Nvariables` represents the number of variables. The `pc` and `fastiamb` labels denote the PC algorithm the Fast IAMB algorithm, respectively.

UPEBNL also achieved lower FDR and SHD (Fig. 3), and the difference became more evident as the number of variables increased. In the PC algorithm scenario with 1,000,000 samples and 500 variables, the FDR of UPEBNL was approximately 0.06, while the FDR values of the bnlearn parallel and non-parallel PC algorithms were approximately 0.5, about 8.3 times higher than that of UPEBNL. This result indicates that UPEBNL reduced false causal dependencies and kept the learned causal knowledge structure closer to the gold standard CBN.

Fig. 4 shows that when the number of variables was small, the runtime of UPEBNL was close to that of the traditional non-parallel algorithm and lower than that of the bnlearn parallel algorithm. As the number of variables increased, the time advantage of UPEBNL became more evident. With 500 variables, the traditional non-parallel Fast IAMB algorithm required approximately 600 minutes, the bnlearn parallel Fast IAMB algorithm required approximately 110 minutes, and UPEBNL required approximately 23 minutes. These results show that UPEBNL improved the efficiency of identifying causal knowledge from large-scale data while maintaining discovery accuracy in high-dimensional settings.

**Fig. 4:**
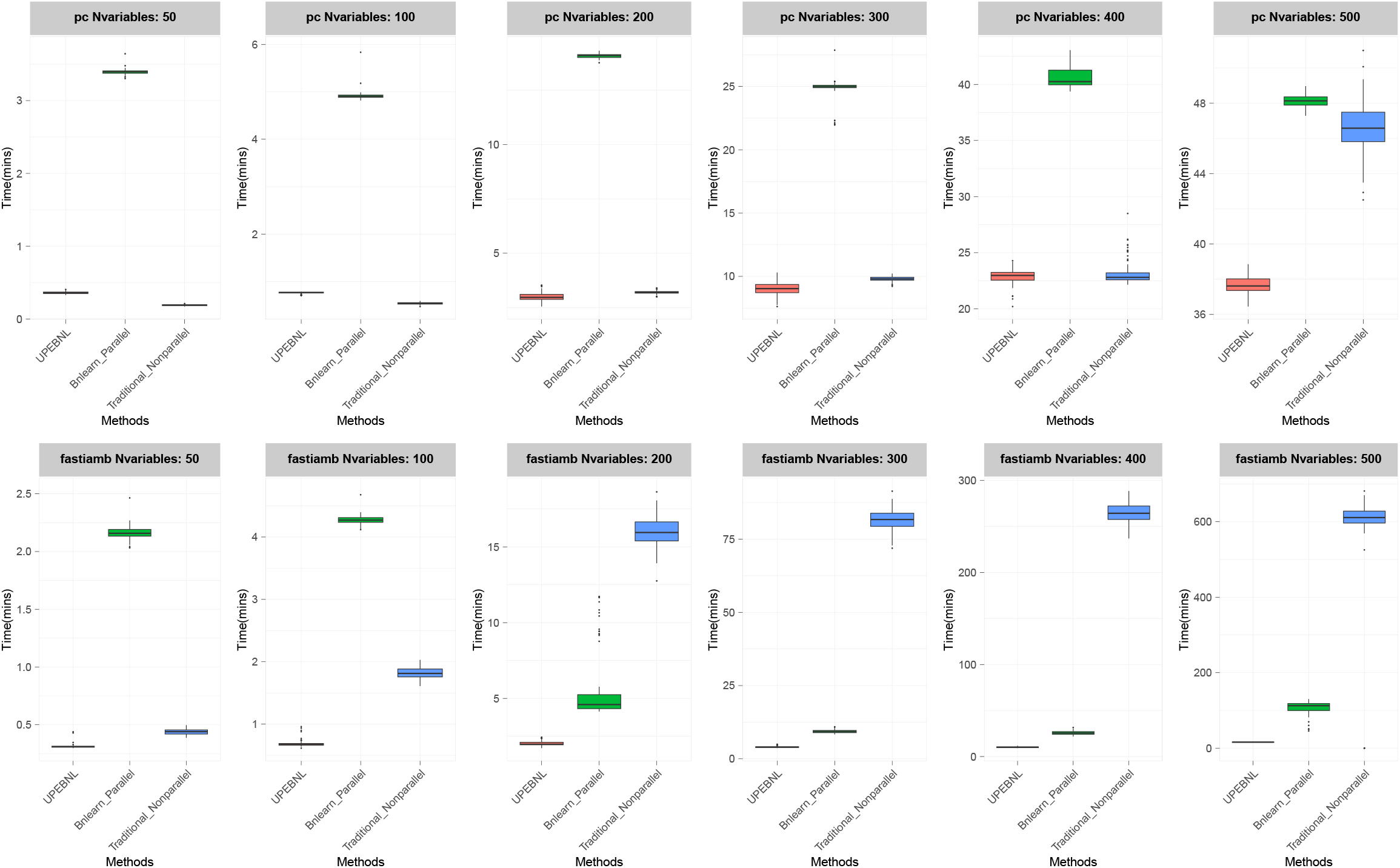
Learning time for structured causal knowledge discovery with 1 million samples and varying numbers of variables. `Nvariables` denotes the number of variables. The `pc` and `fastiamb` labels denote the PC algorithm and the Fast IAMB algorithm, respectively.

### 4.3. Performance under increasing sample size

In the simulations with increasing sample size, the number of variables was fixed at 100 and the network density parameter was set to `prob=NULL`. This setting evaluated the performance of UPEBNL in structured causal knowledge discovery under multi-million-sample conditions. Fig. 5(a)–(c) shows that UPEBNL outperformed the bnlearn parallel and traditional non-parallel algorithms in Precision, Recall, and F1 Score. Fig. 5(d)–(e) shows that UPEBNL also achieved lower FDR and SHD. These results indicate that UPEBNL accurately recovered potential causal dependencies and reduced false dependencies and structural deviations when the sample size increased to the multi-million level.

**Fig. 5:**
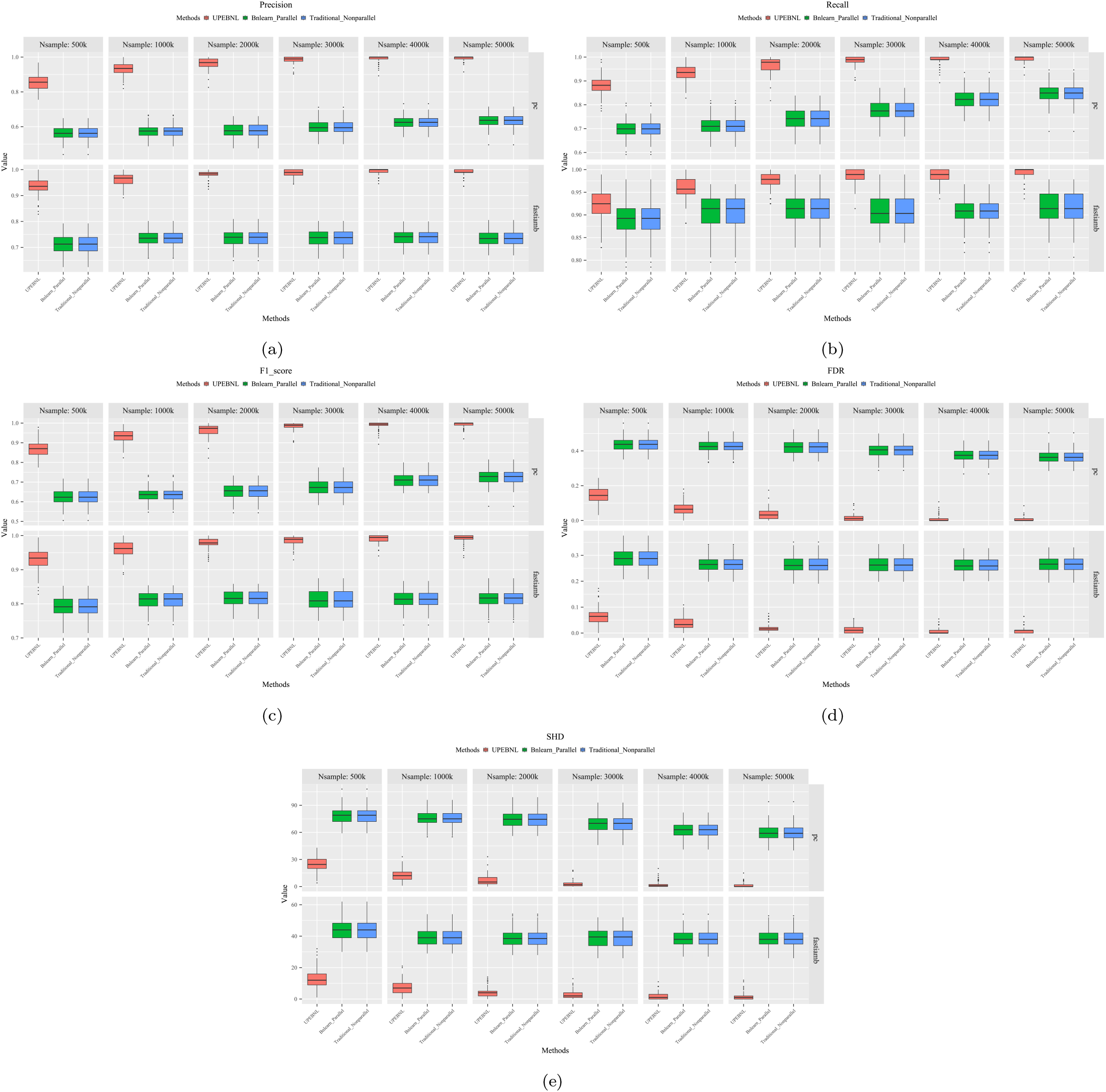
Evaluation of structured causal knowledge discovery with 100 variables and varying sample sizes. The metrics ude (a) Precision, (b) Recall, (c) F1 Score, (d) FDR, and (e) SHD. `Nsample` denotes the number of samples, where the unit “k” rs to thousands.

In terms of learning time (Fig. 6), the runtime of UPEBNL increased only modestly as the sample size grew in sparse networks. For the PC algorithm, UPEBNL required less time than the bnlearn parallel PC algorithm and was close to the traditional non-parallel PC algorithm. For the Fast IAMB algorithm, UPEBNL required less time than the other two implementations. These results indicate that UPEBNL supported structured causal knowledge discovery under multi-million-sample conditions without a marked loss of usability as the sample size increased.

**Fig. 6:**
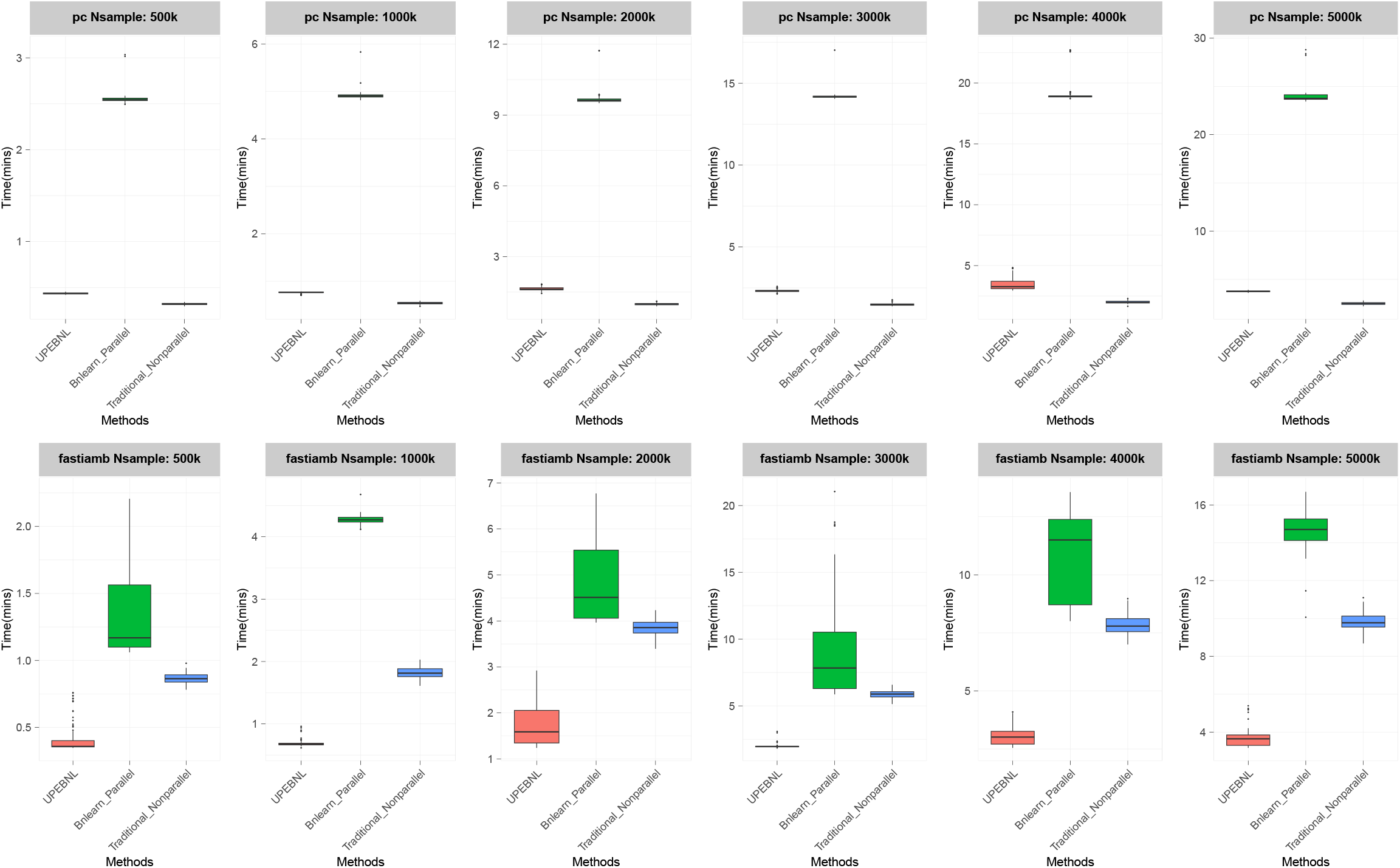
Learning time for structured causal knowledge discovery with 100 variables and 0.5–5 million samples.

### 4.4. Performance under different network densities

In the simulations with varying network density, Fig. 7 shows the results for 1,000,000 samples and 100 variables. UPEBNL achieved higher Precision than the bnlearn parallel and traditional non-parallel algorithms. When the network density parameter `prob`=0.1, the Precision of UPEBNL was 22.5% higher than that of the comparison algorithms. Although the Recall of UPEBNL was slightly lower than that of the other two algorithms, its F1 Score was higher, indicating a better balance between accuracy and completeness.

**Fig. 7:**
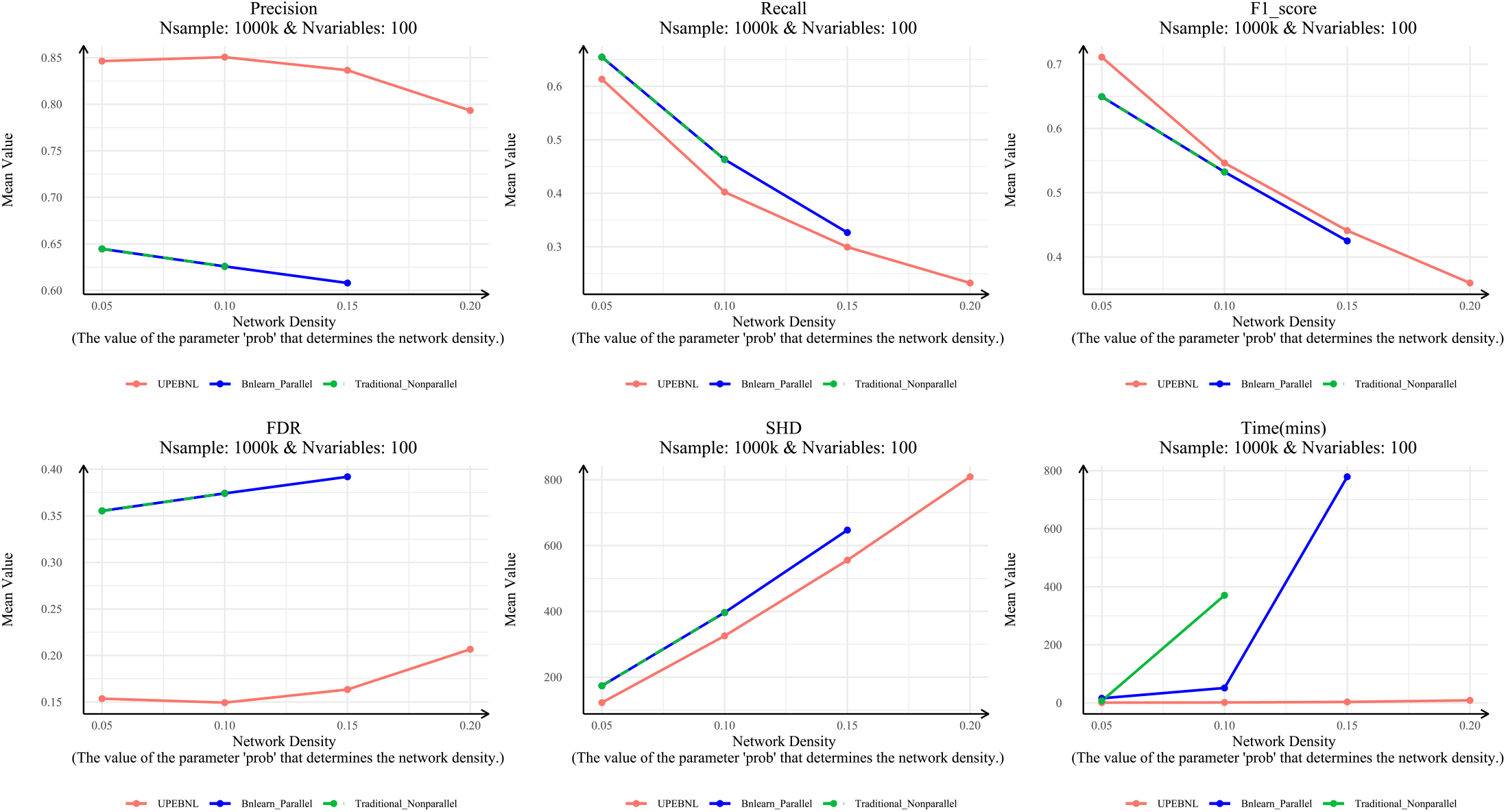
Evaluation of structured causal knowledge discovery under varying levels of causal-dependency complexity, represented by network density, with 1 million samples and 100 variables.

UPEBNL also achieved lower FDR and SHD. As network density increased, the number of candidate causal paths and directional dependencies increased, making false edges and redundant dependencies more likely. Under this condition, UPEBNL still reduced false causal dependencies and maintained smaller structural differences, showing clearer advantages in dense networks.

Fig. 7 also shows that as network density increased, the runtime of the bnlearn parallel and traditional non-parallel algorithms increased rapidly, whereas the runtime of UPEBNL increased more slowly. For example, when `prob` increased from 0.05 to 0.2, the average runtime of UPEBNL increased from 1.04 minutes to 8.84 minutes. When `prob`=0.15, the bnlearn parallel algorithm required 778.92 minutes on average, whereas UPEBNL required 3.52 minutes, corresponding to a 221.28-fold improvement in causal knowledge identification speed. Fig.8 further shows that this gap continued to increase with larger numbers of variables and larger sample sizes. These results indicate that UPEBNL is particularly suitable for discovering complex and dense causal knowledge from data.

**Fig. 8:**
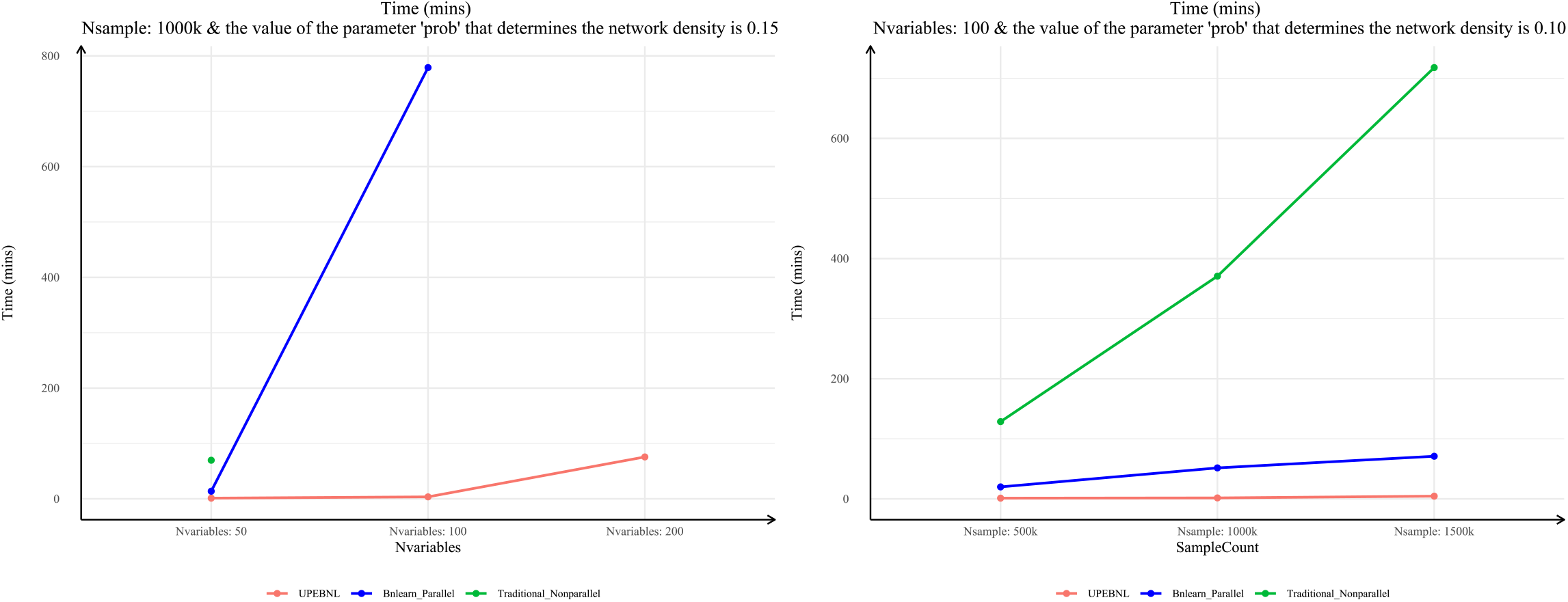
Learning time for structured causal knowledge discovery under dense causal-dependency settings.

Results for other numbers of variables and sample sizes are provided in Supplementary Figs. S1–S4.

### 4.5. UPEBNL with multiple knowledge structure learners

The results (Fig.9) across different knowledge structure learning algorithms show that UPEBNL outperformed the comparison algorithms in Precision, F1 Score, FDR, and SHD, and that the differences increased as the number of variables grew. The Recall of UPEBNL was close to that of the other algorithms. As the number of variables increased, Recall decreased only slightly, by approximately 0.01, and remained close to 1 overall. These results indicate that UPEBNL is applicable not only to constraint-based algorithms, but also to score-based and hybrid knowledge structure learning algorithms, and can identify relatively stable structured knowledge from data across multiple learners. In terms of runtime (Fig.10), UPEBNL also required less time than the comparison algorithms.

**Fig. 9:**
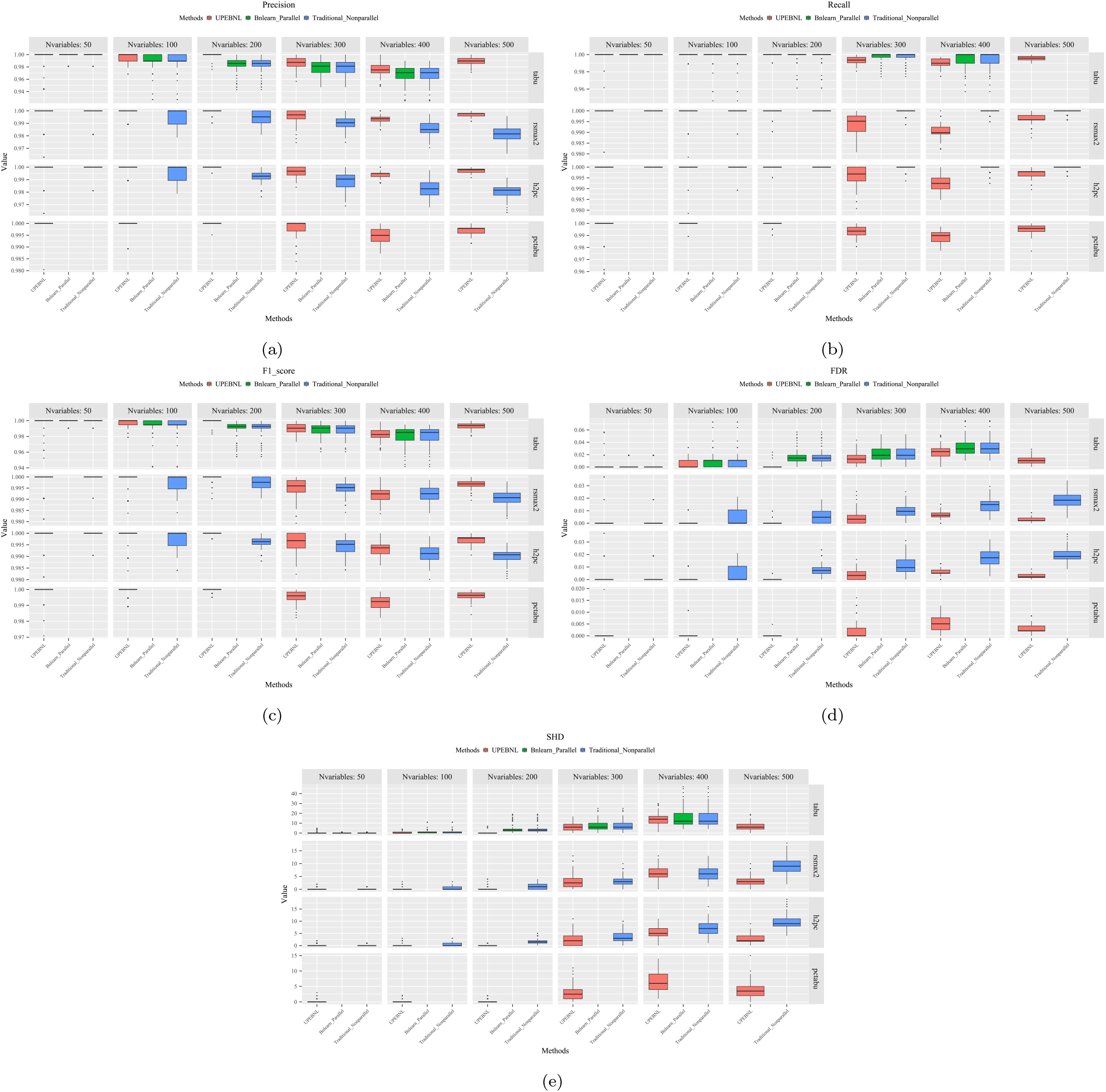
Evaluation of structured knowledge discovery with score-based and hybrid learners under 1 million samples.

**Fig. 10:**
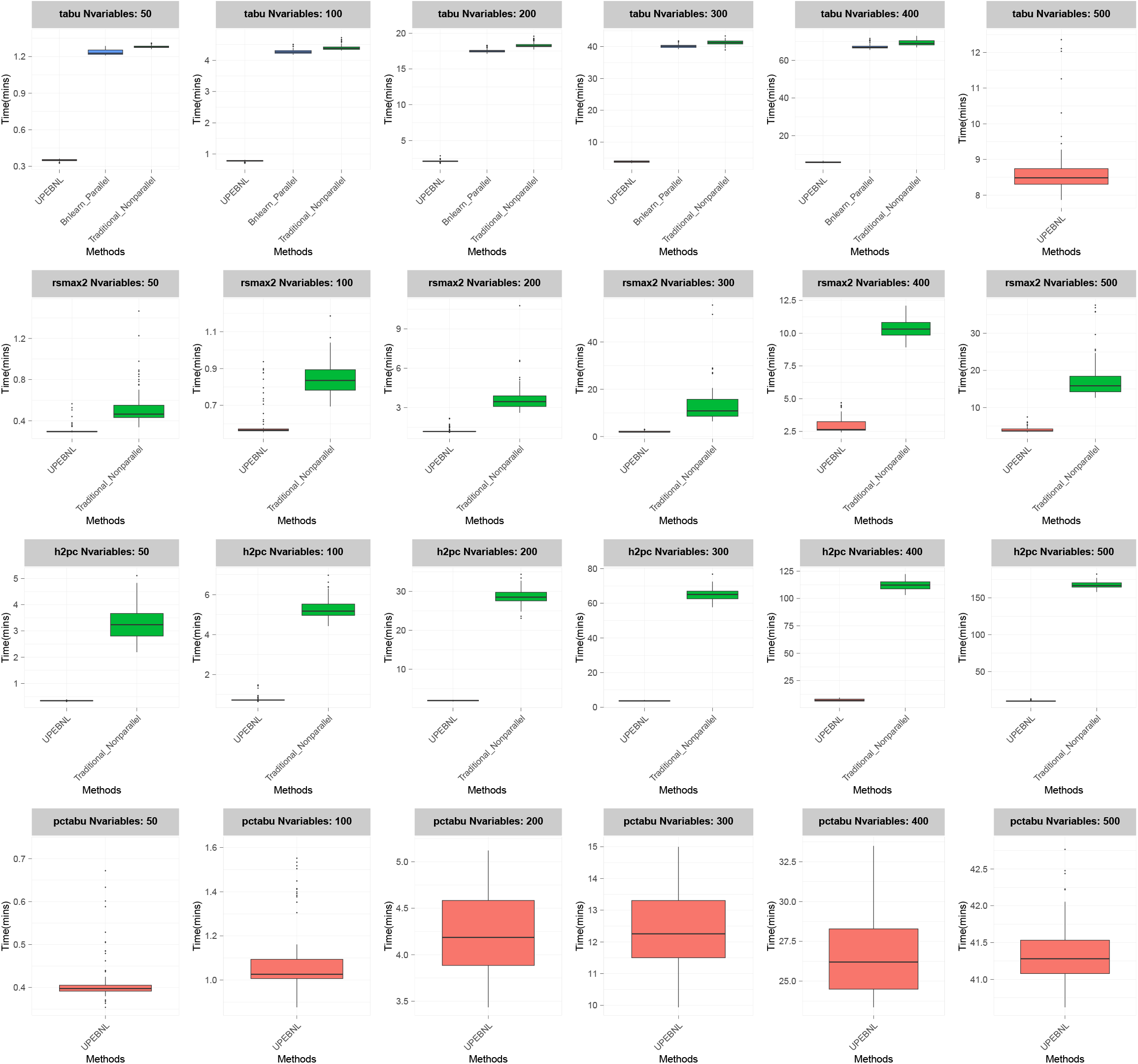
Learning time for structured knowledge discovery with score-based and hybrid learners under 1 million samples.

In summary, these results show that the advantage of UPEBNL is not limited to reducing learning time. It also maintains the accuracy and usability of causal knowledge discovery at larger-scale data.

## 5. EHR-based cancer prediction application

Conventional cancer screening methods, such as endoscopy [38], allow direct visualization of lesions and histological confirmation, and therefore have clear clinical value for cancer screening and early diagnosis. However, these methods are invasive, costly, and dependent on specialized equipment. In resource-limited settings or large-scale population screening, using endoscopy as the initial screening tool may be constrained by limited accessibility and low adherence. A more practical strategy is to first stratify individuals using low-cost and readily available data, so that those at higher risk can be prioritized for further specialist evaluation.

EHR data provide a practical basis for such prescreening. They continuously record demographic characteristics, prior diseases, clinical diagnoses, medication use, care-seeking behavior, and health service utilization in real-world clinical settings. These longitudinal records can reflect changes in health status, accumulation of comorbidities, and evolution of potential risk factors over time. EHR-based risk prediction can therefore serve as a low-cost and scalable prescreening tool for identifying high-risk individuals within the general population or health-system-covered populations, thereby improving the efficient use of downstream screening resources.

Motivated by this rationale, we used UPEBNL to learn cancer-specific CBNs from large-scale EHR data. The learned networks were used to characterize dependency paths among risk factors, clinical indicators, and cancer outcomes related to esophageal and colorectal cancer, and to support individual risk prediction through CBN-based probabilistic reasoning. Unlike black-box models that mainly provide risk scores, CBNs offer an explicit structural representation that can help interpret risk propagation pathways and key contributing factors.

### 5.1. Data source

We used data from the Cheeloo LEAD database [39], which integrates information from 5,152,597 individuals in 39 urban areas of Shandong Province, China. Each individual can be linked to comprehensive data across 4,909 medical institutions through an encrypted unique ID. The Cheeloo LEAD database retrospectively collected EHR data from 2009 to 2024. In this study, we selected records from January 1, 2015, to October 31, 2017, and included individuals aged 40 years and above. The data were randomly divided into training and validation sets in a 7:3 ratio.

### 5.2. Learned causal knowledge structures for cancer prescreening

Univariate logistic regression models and Incremental Feature Selection (IFS) [40] were used to select 71 variables from 5,519 potential predictors for constructing the cancer-specific CBNs.

The CBNs were learned on the training set using the PC algorithm within UPEBNL. To improve structural stability, we used a bootstrap strategy [41]. The procedure performed 50 iterations of network learning and retained an edge in the final network when the edge appeared in at least 70% of the learned networks. CBN parameters were then estimated using Bayesian estimation [42]. The risks of esophageal cancer and colorectal cancer were predicted through CBN uncertainty reasoning [43, 44, 45].

The learned CBNs for esophageal cancer and colorectal cancer are shown in Fig.11. These structures organize EHR-derived variables into interpretable dependency paths related to cancer risk. In the esophageal cancer CBN, age, sex, esophageal diseases, and stomach and duodenal diseases were directly connected to future esophageal cancer occurrence. These variables can be interpreted as direct risk factors for esophageal cancer prescreening in the learned CBN. In the colorectal cancer CBN, age, sex, and intestinal diseases were directly connected to future colorectal cancer occurrence. These direct connections provide interpretable evidence paths for cancer prescreening, while other EHR variables may still contribute through indirect dependency paths.

**Fig. 11:**
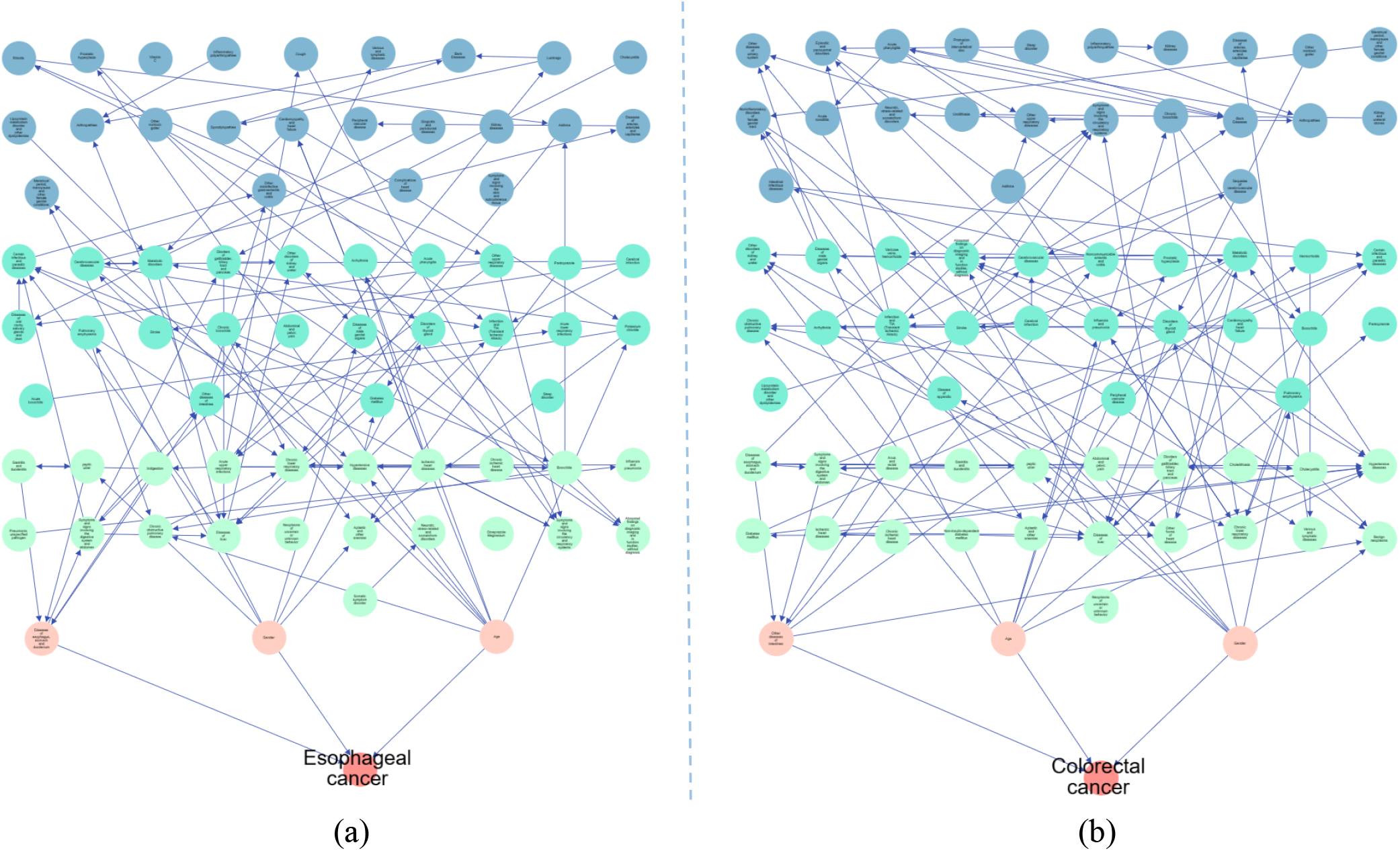
Learned CBNs for esophageal cancer and colorectal cancer prescreening. The left panel shows the CBN for esophageal cancer, and the right panel shows the CBN for colorectal cancer. The figure can be enlarged on the webpage (https://otherrun2.aiself.net/) for a better view.

### 5.3. Application results

The prescreening models were evaluated using calibration [46], discrimination [47], and clinical utility [48]. Calibration measures agreement between predicted risks and observed outcomes. Discrimination was evaluated using the area under the receiver operating characteristic curve (AUC). Clinical utility was evaluated by decision curve analysis across threshold probabilities.

For esophageal cancer, the prescreening model based on the learned causal knowledge structure achieved an AUC of 0.827 (95% CI: 0.8174–0.836) in the training set, as shown in Fig.12. In the validation set, the AUC was 0.8171 (95% CI: 0.8022–0.8319), as shown in Fig.13. The calibration curves showed good agreement between predicted and observed risks in both sets. Decision curve analysis also indicated clinical utility across relevant threshold probabilities.

**Fig. 12:**
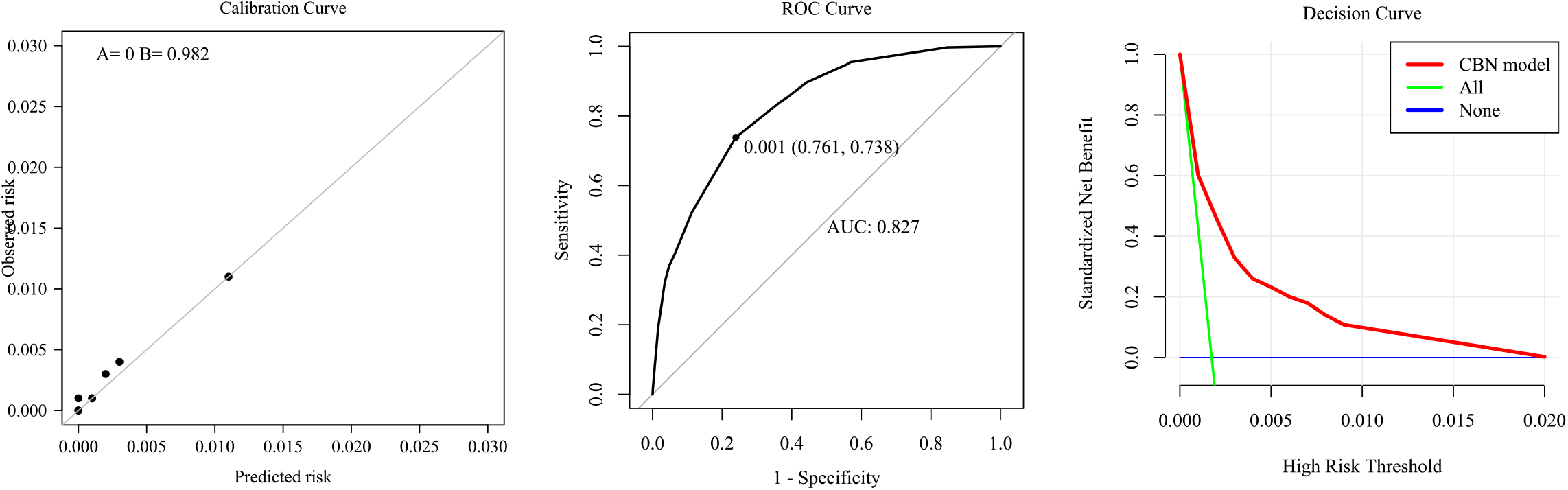
Model evaluation of esophageal cancer in the training set. The left panel shows the calibration curve, with ‘A’ representing the intercept and ‘B’ the slope. The middle panel presents the ROC curve. The right panel shows the decision curve.

**Fig. 13:**
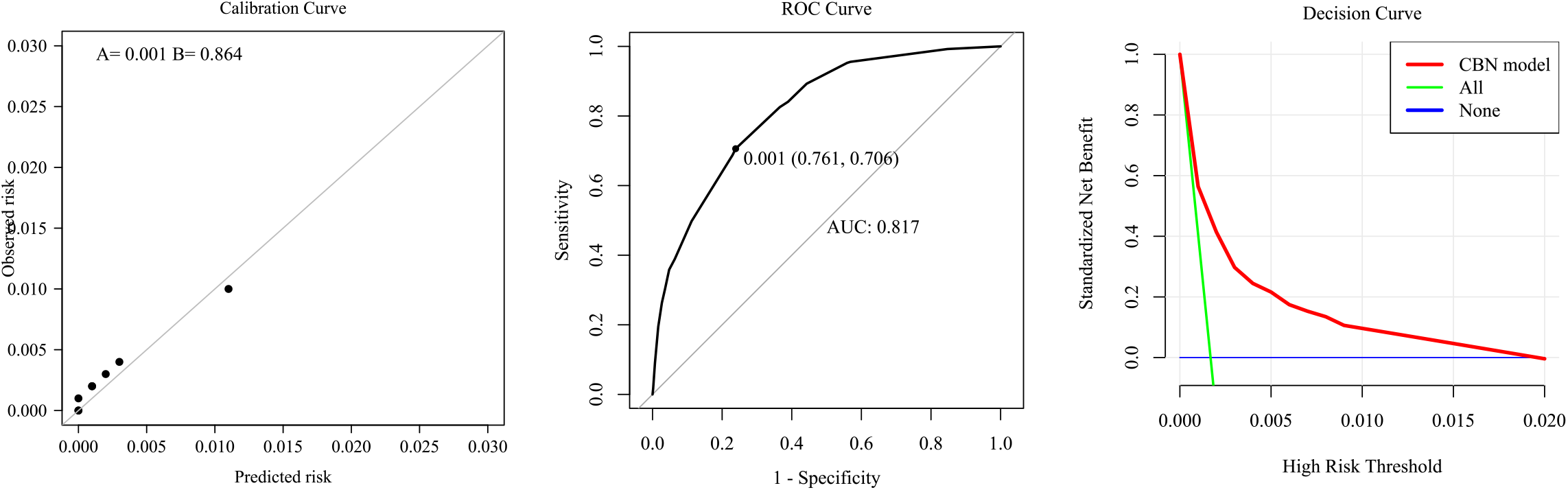
Model evaluation of esophageal cancer in the validation set. The left panel shows the calibration curve, with ‘A’ representing the intercept and ‘B’ the slope. The middle panel presents the ROC curve. The right panel shows the decision curve.

For colorectal cancer, the prescreening model achieved an AUC of 0.776 (95% CI: 0.766–0.786) in the training set, as shown in Fig. 14, and 0.784 (95% CI: 0.769–0.799) in the validation set, as shown in Fig.15. The colorectal cancer model also showed good calibration and favorable net clinical benefit.

**Fig. 14:**
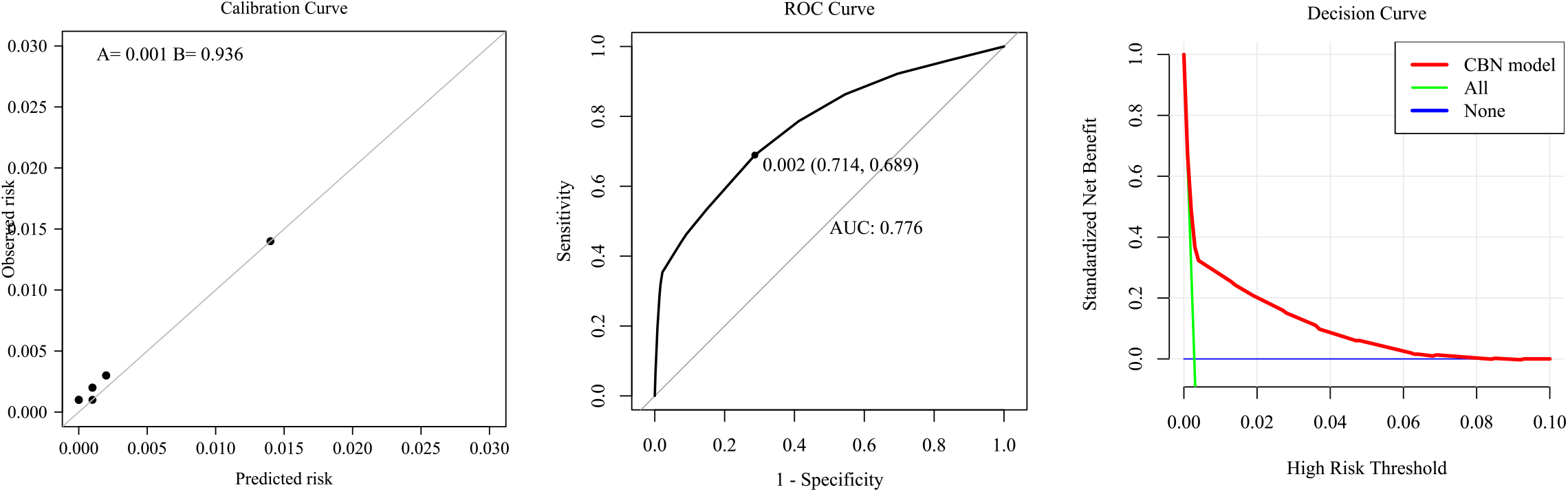
Model evaluation of colorectal cancer in the training set. The left panel shows the calibration curve, with ‘A’ representing the intercept and ‘B’ the slope. The middle panel presents the ROC curve. The right panel shows the decision curve.

**Fig. 15:**
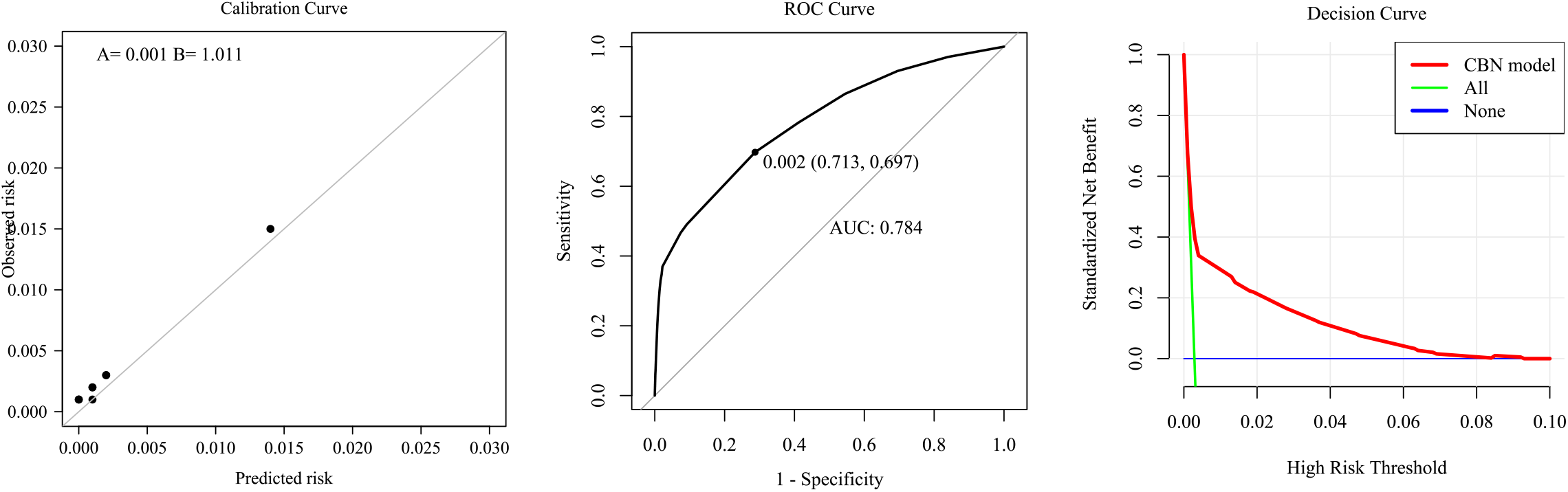
Model evaluation of colorectal cancer in the validation set. The left panel shows the calibration curve, with ‘A’ representing the intercept and ‘B’ the slope. The middle panel presents the ROC curve. The right panel shows the decision curve.

These results show that UPEBNL can learn cancer-related structured causal knowledge from large-scale EHR data and use this knowledge to support interpretable cancer risk prescreening. The learned CBNs provided visible evidence paths for esophageal and colorectal cancer prediction, while the resulting prescreening models showed good discrimination, calibration, and clinical utility.

## 6. Discussion and Conclusion

This study focuses on scalable causal knowledge discovery from large-scale observational data. The proposed UPEBNL framework learns CBN structures through adaptive data partitioning, quality-aware aggregation, and global DAG restoration, making CBN-based causal knowledge learning feasible for large-scale data.

Simulation results show that UPEBNL improves structural recovery and reduces computation time in high-dimensional and million-sample settings. As the number of variables increased, UPEBNL maintained higher precision and F1 scores, as well as lower FDR and SHD, than the compared parallel and non-parallel strategies. This advantage largely arises from the divide-and-conquer strategy adopted by UPEBNL. Through this strategy, UPEBNL enables complementary correction across subsets and substantially reduces the risk of overfitting that may occur when learning from a single dataset, thereby improving the overall accuracy and stability of the network. In the future, the aggregation idea of UPEBNL can be extended to causal knowledge learning from multi-center data, where data from each center can be treated as a data slice. This would preserve data security while allowing more comprehensive and stable structured causal knowledge to be obtained.

The most evident advantage of UPEBNL appeared in dense networks. As network density increased, the computation time of the bnlearn parallel algorithm and the traditional non-parallel algorithm increased sharply, whereas UPEBNL showed a more moderate increase. In a setting with 1 million samples and 100 variables, UPEBNL required only 3.52 minutes, while the bnlearn parallel algorithm required 778.92 minutes, corresponding to a 221.28-fold speed improvement. This result is important because real-world systems often contain complex dependencies, and their network density is usually unknown before structure learning.

The EHR application further illustrates the practical value of the learned causal knowledge for real-world decision support. In the esophageal and colorectal cancer tasks, the CBNs captured explicit dependency structures linking EHR-derived variables to cancer outcomes. These structures supported risk prediction models that achieved useful discrimination, calibration, and clinical utility in both the training and validation sets. These findings indicate that learned CBNs can serve as interpretable prescreening tools for identifying individuals who may require further evaluation before invasive examination.

From the perspective of knowledge-based systems, the contribution of UPEBNL is not limited to improving computational efficiency. The framework provides a scalable route from large-scale data to explicit DAG-based causal knowledge. The final CBN can be queried with evidence and used for probabilistic reasoning.

The learned CBNs may also serve as external causal scaffolds for decision systems based on large language models (LLMs). By encoding evidence propagation paths, CBNs can provide a structured basis for organizing information during model reasoning. Such structures may help LLMs rely less on spurious associations and support more mechanism-aware decision-making.

Several limitations remain. The current experiments used discrete data, and further simulation evaluation on continuous data is needed. The EHR application was conducted using one large regional database, so external validation is still required before clinical deployment.

In conclusion, UPEBNL provides a scalable framework for learning structured causal knowledge from large-scale data. By combining adaptive partitioning, weighted ensemble learning, and global DAG restoration, the method improves the feasibility of CBN-based structured causal knowledge discovery in high-dimensional, large-sample, and complex data settings. The EHR cancer prescreening study further demonstrates its potential for interpretable real-world decision support.

## Ethics statement

This study was approved by the Institutional Review Board of the School of Public Health, Shandong University, China (approval number: LL20240308). The requirement for informed consent was waived by the Institutional Review Board because the study used anonymized electronic health record data.

## Declaration of generative AI and AI-assisted technologies in the manuscript preparation process

During the preparation of this work, the authors used AI to review grammar and improve clarity in the text. After using this tool, the authors reviewed and edited the content as needed and take full responsibility for the content of the published article.

## CRediT authorship contribution statement

Shuaijie Zhang: Methodology, Validation, Formal analysis, Data curation, Visualization, Writing - original draft, Writing - review and editing. Fuzhong Xue: Conceptualization, Methodology, Supervision, Funding acquisition, Writing - review and editing.

## Data availability

The simulation datasets were generated according to the procedures described in the paper. The real-world EHR data are not publicly available because they contain sensitive health information and are subject to institutional data governance restrictions. Access may be granted by the corresponding author upon reasonable request and completion of a data security agreement.

## Acknowledgment

This study was supported by the Key Program of the National Natural Science Foundation of China (Grant No. 82330108); the China Postdoctoral Science Foundation (grant 2026M790787); the National Key Research and Development Program of China (grant 2020YFC2003500); and the Research and Application Demonstration of Key Technologies and Products for Trustworthy Artificial Intelligence Agents in Health Big Data (grant 202534084).

## Supplementary material

Additional pseudocode and simulation results are provided in the Supplementary Material.

## References

[1] A. J. Thirunavukarasu, D. S. J. Ting, K. Elangovan, L. Gutierrez, T. F. Tan, D. S. W. Ting, Large language models in medicine, Nature Medicine 29 (2023) 1930–1940. doi:10.1038/s41591-023-02448-8.

[2] V. Hassija, V. Chamola, A. Mahapatra, A. Singal, D. Goel, K. Huang, S. Scardapane, I. Spinelli, M. Mahmud, A. Hussain, Interpreting black-box models: A review on explainable artificial intelligence, Cognitive Computation 16 (2024) 45–74. doi:10.1007/s12559-023-10179-8.

[3] C. Kim, S. U. Gadgil, S.-I. Lee, Transparency of medical artificial intelligence systems, Nat. Rev. Bioeng. 4 (2026) 11–29. doi:10.1038/s44222-025-00363-w.

[4] S. Miret, N. M. A. Krishnan, Enabling large language models for real-world materials discovery, Nature Machine Intelligence 7 (2025) 991–998. doi:10.1038/s42256-025-01058-y.

[5] S. Feuerriegel, D. Frauen, V. Melnychuk, J. Schweisthal, K. Hess, A. Curth, S. Bauer, N. Kilbertus, S. Kohane, M. van der Schaar, Causal machine learning for predicting treatment outcomes, Nature Medicine 30 (2024) 958–968. doi:10.1038/s41591-024-02902-1.

[6] I. Olier, Y. Zhan, X. Liang, V. Volovici, et al., Causal inference and observational data, BMC Medical Research Methodology 23 (2023) 227. doi:10.1186/s12874-023-02058-5.

[7] K. Lagemann, C. Lagemann, B. Taschler, S. Mukherjee, Deep learning of causal structures in high dimensions under data limitations, Nature Machine Intelligence 5 (2023) 1306–1316. doi:10.1038/s42256-023-00744-z.

[8] N. K. Kitson, A. C. Constantinou, Z. Guo, Y. Liu, K. Chobtham, A survey of Bayesian Network structure learning, Artif. Intell. Rev. 56 (2023) 8721–8814. doi:10.1007/s10462-022-10351-w.

[9] J. Pearl, Causality: Models, Reasoning, and Inference, 2nd Edition, Cambridge University Press, Cambridge, 2009.

[10] J. M. Ordovas, D. Rios-Insua, A. Santos-Lozano, A. Lucia, A. Torres, A. Kosgodagan, J. M. Camacho, A bayesian network model for predicting cardiovascular risk, Computer Methods and Programs in Biomedicine 231 (2023) 107405. doi:10.1016/j.cmpb.2023.107405.

[11] P. Spirtes, C. Glymour, An algorithm for fast recovery of sparse causal graphs, Soc. Sci. Comput. Rev. 9 (1) (1991) 62–72. doi:10.1177/089443939100900106.

[12] M. Kalisch, P. Bühlmann, Estimating high-dimensional directed acyclic graphs with the PC-algorithm, J. Mach. Learn. Res. 8 (2007) 613–636.

[13] A. Zanga, E. Ozkirimli, F. Stella, A survey on causal discovery: Theory and practice, Int. J. Approx. Reason. 151 (2022) 101–129. doi:10.1016/j.ijar.2022.09.004.

[14] R. Arone, H. Caetano, C. Maciel, Quantifying causal effects to enhance explainability in Causal Bayesian Networks, Knowl.-Based Syst. 338 (2026) 115495. doi:10.1016/j.knosys.2026.115495.

[15] J. Pearl, Probabilistic Reasoning in Intelligent Systems: Networks of Plausible Inference, Morgan Kaufmann, San Mateo, CA, 1988.

[16] C. Rudin, Stop explaining black box machine learning models for high stakes decisions and use interpretable models instead, Nat. Mach. Intell. 1 (5) (2019) 206–215. doi:10.1038/s42256-019-0048-x.

[17] P. Spirtes, C. Glymour, R. Scheines, Causation, Prediction, and Search, 2nd Edition, MIT Press, Cambridge, MA, 2001.

[18] M. Scutari, Bayesian Network constraint-based structure learning algorithms: Parallel and optimized implementations in the bnlearn R package, J. Stat. Softw. 77 (2) (2017) 1–20. doi:10.18637/jss.v077.i02.

[19] S. Yaramakala, D. Margaritis, Speculative Markov blanket discovery for optimal feature selection, in: Proceedings of the Fifth IEEE International Conference on Data Mining, IEEE Computer Society, 2005, pp. 809–812. doi:10.1109/ICDM.2005.134.

[20] G. Schwarz, Estimating the dimension of a model, Ann. Stat. 6 (2) (1978) 461–464. doi:10.1214/aos/1176344136.

[21] D. M. Chickering, Learning equivalence classes of Bayesian-network structures, J. Mach. Learn. Res. 2 (2002) 445–498.

[22] I. Tsamardinos, L. E. Brown, C. F. Aliferis, The max-min hill-climbing bayesian network structure learning algorithm, Machine learning 65 (2006) 31–78.

[23] T. D. Le, T. Hoang, J. Li, L. Liu, H. Liu, S. Hu, A fast PC algorithm for high dimensional causal discovery with multi-core PCs, IEEE/ACM Trans. Comput. Biol. Bioinform. 16 (5) (2019) 1483–1495. doi:10.1109/TCBB.2016.2591526.

[24] D. Colombo, M. H. Maathuis, Order-independent constraint-based causal structure learning, J. Mach. Learn. Res. 15 (2014) 3921–3962.

[25] Y. Tang, J. Wang, M. Nguyen, I. Altintas, PENbayes: A multi-layered ensemble approach for learning Bayesian network structure from big data, Sensors 19 (20) (2019) 4400. doi:10.3390/s19204400.

[26] T. Huang, Y. Zhou, Partitioned hybrid learning of Bayesian network structures, Mach. Learn. 111 (2022) 1695–1738. doi:10.1007/s10994-022-06145-4.

[27] A. Srivastava, S. P. Chockalingam, S. Aluru, A parallel framework for constraint-based Bayesian network learning via Markov blanket discovery, IEEE Trans. Parallel Distrib. Syst. 34 (6) (2023) 1699–1715. doi:10.1109/TPDS.2023.3244135.

[28] I. Tsamardinos, C. F. Aliferis, A. R. Statnikov, Algorithms for large scale Markov blanket discovery, in: Proceedings of the Sixteenth International Florida Artificial Intelligence Research Society Conference, AAAI Press, 2003, pp. 376–381.

[29] A. L. Madsen, F. Jensen, A. Salmerón, H. Langseth, T. D. Nielsen, A parallel algorithm for Bayesian network structure learning from large data sets, Knowl. - Based Syst. 117 (2017) 46–55. doi:10.1016/j.knosys.2016.07.031.

[30] J. Yang, J. Jiang, Z. Wen, A. Mian, Parallel and distributed Bayesian network structure learning, IEEE Trans. Parallel Distrib. Syst. 35 (4) (2024) 517–530. doi:10.1109/TPDS.2023.3326832.

[31] W. Buntine, Theory refinement on bayesian networks, in: Uncertainty proceedings 1991, Elsevier, 1991, pp. 52–60.

[32] D. Heckerman, D. Geiger, D. M. Chickering, Learning bayesian networks: The combination of knowledge and statistical data, Machine learning 20 (1995) 197–243.

[33] J. Pearl, M. Glymour, N. P. Jewell, Causal inference in statistics: A primer, John Wiley & Sons, 2016.

[34] J. Yang, J. Jiang, Z. Wen, A. Mian, Parallel and distributed bayesian network structure learning, IEEE Transactions on Parallel and Distributed Systems 35 (4) (2024) 517–530. doi:10.1109/TPDS.2023.3326832.

[35] D. M. Chickering, A transformational characterization of equivalent bayesian network structures, arXiv preprint arXiv:1302.4938 (2013).

[36] M. Scutari, C. E. Graafland, J. M. Gutiérrez, Who learns better bayesian network structures: Accuracy and speed of structure learning algorithms, International Journal of Approximate Reasoning 115 (2019) 235–253.

[37] F. Glover, Tabu search: A tutorial, Interfaces 20 (4) (1990) 74–94.

[38] J. A. Kim, P. M. Shah, Screening and prevention strategies and endoscopic management of early esophageal cancer, Chinese clinical oncology 6 (5) (2017) 50–50.

[39] S. Zhang, Q. Wang, X. Hu, et al., Interpretable machine learning model for digital lung cancer prescreening in chinese populations with missing data, npj Digital Medicine 7 (2024) 327. doi: 10.1038/s41746-024-01309-z.

[40] H. Liu, R. Setiono, Incremental feature selection, Applied Intelligence 9 (3) (1998) 217–230. doi: 10.1023/A:1008363719778.

[41] N. Friedman, M. Goldszmidt, A. J. Wyner, Data analysis with bayesian networks: A bootstrap approach, in: UAI ‘99: Proceedings of the Fifteenth Conference on Uncertainty in Artificial Intelligence, Morgan Kaufmann, 1999, pp. 196–205.

[42] Z. Ji, Q. Xia, G. Meng, A review of parameter learning methods in bayesian network, in: Advanced Intelligent Computing Theories and Applications: 11th International Conference, ICIC 2015, Fuzhou, China, August 20-23, 2015. Proceedings, Part III 11, Springer, 2015, pp. 3–12.

[43] Y. Peng, S. Zhang, R. Pan, Bayesian network reasoning with uncertain evidences, International Journal of Uncertainty, Fuzziness and Knowledge-Based Systems 18 (05) (2010) 539–564.

[44] R. D. Shachter, M. A. Peot, Simulation approaches to general probabilistic inference on belief networks, in: Machine intelligence and pattern recognition, Vol. 10, Elsevier, 1990, pp. 221–231.

[45] R. Fung, K.-C. Chang, Weighing and integrating evidence for stochastic simulation in bayesian networks, in: Machine intelligence and pattern recognition, Vol. 10, Elsevier, 1990, pp. 209–219.

[46] Y. Huang, W. Li, F. Macheret, R. A. Gabriel, L. Ohno-Machado, A tutorial on calibration measurements and calibration models for clinical prediction models, Journal of the American Medical Informatics Association 27 (4) (2020) 621–633.

[47] E. W. Steyerberg, Y. Vergouwe, Towards better clinical prediction models: seven steps for development and an abcd for validation, European heart journal 35 (29) (2014) 1925–1931.

[48] A. J. Vickers, F. Holland, Decision curve analysis to evaluate the clinical benefit of prediction models, The Spine Journal 21 (10) (2021) 1643–1648.

