## Supplementary Material for "Scalable Causal-Interpretable Machine Learning for Cancer Prescreening Using Electronic Health Records"

### S1. Multi-Learner Structured Causal Knowledge Learning with UPEBNL

**Algorithm S1** Determining appropriate learning size with prior knowledge ($ALS\_with\_PK$) for multiple knowledge structure learners

**Input:**
$D$: Dataset
$\beta, \epsilon$: Thresholds
$mstep$: Maximum loop steps
$InitialSize$: Initial data slice size
$white\_list$: Prior knowledge (must-include edges)
$black\_list$: Prior knowledge (must-not-include edges)
$stru\_meth\_num$: The number of structure learning algorithms used
$algor$: The knowledge structure learners are denoted as $\{algor_1, algor_2, \ldots\}$.

**Output:**
$ALS\_with\_PK$: Appropriate Learning Size with Prior Knowledge

```
bestAMBS ← 1, bestES ← −1, step ← 1, sliceSize ← InitialSize        ▷ Initialize parameters.
while step < mstep do
    Sample sliceSize rows from D to form data_slice
    if stru_meth_num = 1 then
        Learn BN structure CBN_{data_slice}: algor(data_slice, white_list, black_list)
    else if stru_meth_num ≥ 2 then
        Learn CBN structure CBN_{data_slice} using the local ensemble algorithm (Algorithm S2): local_ensemble(data_slice, white_list, black_list)
    end if
    AMBS ← Average Markov Blanket Size (AMBS) of BN_{data_slice}
    Score(CBN_{data_slice}, data_slice) = BDeu(CBN_{data_slice}, data_slice)
    ES(CBN_{data_slice}) = Score(CBN_{data_slice}, data_slice) / (Nsamples × Medges)
    if (|AMBS − bestAMBS| > bestAMBS × β) or (|ES − bestES| > bestES × ϵ) then
        bestAMBS ← AMBS
        bestES ← ES
        sliceSize ← sliceSize × 2
    end if
    step ← step + 1
end while
ALS_with_PK ← sliceSize
return ALS_with_PK
```

*Corresponding author

**Algorithm S2** UPEBNL for integrating multiple knowledge structure learners: local ensemble

**Input:**
$data\_slice$: Data slice
$stru\_meth\_num$: The number of structure learning algorithms used.
$white\_list$: Prior knowledge (must-include edges)
$black\_list$: Prior knowledge (must-not-include edges)
$algor$: The knowledge structure learners are denoted as $\{algor_1, algor_2, \ldots\}$.
$T$: Structure ensemble threshold factor
**Output:**
$CBN_{local}$: Local causal knowledge structure learned from a single data slice.
$AM(CBN_{local})$: The adjacency matrices of the CBN learned from a single data slice.
$ES(CBN_{local})$: Edge strength

**if** stru_meth_num = 1 **then**
  $CBN_{local} \leftarrow algor(data\_slice, white\_list, black\_list)$
  Score($CBN_{local}, data\_slice$) = BDeu($CBN_{local}, data\_slice$)
  $ES(CBN_{local}) = \frac{Score(CBN_{local}, data\_slice)}{N_{samples} \times M_{edges}}$
  $AM(CBN_{local})$ ▷ The adjacency matrix of $CBN_{local}$.
**else**
  **if** stru_meth_num $\geq 2$ **then**
    $AM \leftarrow \emptyset$ ▷ A list storing adjacency matrices.
    $ES \leftarrow \emptyset$ ▷ A vector storing edge weights.
    **for** $i = 1$ to $stru\_meth\_num$ **do**
      $BN_{local\_algor_i} \leftarrow algor_i(data\_slice, white\_list, black\_list)$
      Score($CBN_{local\_algor_i}, data\_slice$) = BDeu($CBN_{local\_algor_i}, data\_slice$)
      $ES[i] = \frac{Score(CBN_{local\_algor_i}, data\_slice)}{N_{samples} \times M_{edges_{CBN_{local\_algor_i}}}}$
      $AM[i] = AM(CBN_{local\_algor_i})$ ▷ The adjacency matrix of $CBN_{local\_algor_i}$.
    **end for**
    **for** $i = 1$ to $stru\_meth\_num$ **do**
      $W(CBN_{local\_algor_i}) = \frac{ES(i)}{\sum_{i=1}^{stru\_meth\_num} ES(i)}$
      $WAM_{CBN_{local\_algor_i}} = AM[i] \times W(CBN_{local\_algor_i})$, $i = \{1, 2, \ldots, stru\_meth\_num\}$
    **end for**
    $FWAM = \sum_{i=1}^{stru\_meth\_num} WAM_{CBN_{local\_algor_i}}$
    $\gamma = T \times \min(W(CBN_{local\_algor_i}))$ ▷ Set the structure ensemble threshold.
    **for** each $i, j$ in $FWAM$ **do**
      **if** $FWAM[i, j] \geq \gamma$ **then**
        $EAM[i, j] = 1$ ▷ Ensemble adjacency matrix.
      **else**
        $EAM[i, j] = 0$
      **end if**
    **end for**
    **if** bidirectional edges exist **then**
      $node\_list \leftarrow nodes(G)$ ▷ Initialize node list and combinations.
      $allarcs \leftarrow$ all possible combinations of any two nodes in the $node\_list$
      $WL \leftarrow$ directed arcs($G$) as a dataframe ▷ Retrieve directed and undirected arcs from $G$.
      $complete\_arc \leftarrow$ combine WL and undirected arcs($G$)
      $BL \leftarrow$ edges in $allarcs$ not in $complete\_arc$ ▷ Identify blacklisted edges.
      $result\_DAG \leftarrow minBIC(Data, whitelist = WL, blacklist = BL)$, update $EAM$
    **end if**
    $AM(CBN_{local}) = EAM$ ▷ Set the final ensembled network.
    $ES(CBN_{local}) = \text{mean}(ES)$
  **end if**
**end if**
**return** $CBN_{local}, AM(CBN_{local}), ES(CBN_{local})$

**Algorithm S3** UPEBNL for integrating multiple knowledge structure learners: global ensemble

**Input:**

$Nd$: The number of data slices

$\mathbf{AM}(CBN_{local}) = \{AM(CBN_{local_1}), AM(CBN_{local_2}), \ldots, AM(CBN_{local_{Nd}})\}$: The adjacency matrices of the CBN learned from each data slice.

$\mathbf{ES}(CBN_{local}) = \{ES(CBN_{local_1}), ES(CBN_{local_2}), \ldots, ES(CBN_{local_{Nd}})\}$: The edge strength of the CBN learned from each data slice.

$T$: Structure ensemble threshold factor

**Output:** $CBN_{global}$

**for** $i = 1$ **to** $Nd$ **do**
    $W(CBN_{local_i}) = \frac{ES(CBN_{local_i})}{\sum_{i=1}^{Nd} ES(CBN_{local_i})}$
    $WAM_{CBN_{local_i}} = AM[i] \times W(CBN_{local_i}), i = \{1, 2, \ldots, Nd\}$
**end for**
$FWAM = \sum_{i=1}^{Nd} WAM_{CBN_{local_i}}$
$\gamma = T \times \min(W(CBN_{local_i}))$ ▷ Set the structure ensemble threshold.
**for** each element $[i, j]$ in $FWAM$ **do**
    **if** $FWAM[i, j] \geq \gamma$ **then**
        Ensemble adjacency matrix $EAM[i, j] = 1$
    **else**
        $EAM[i, j] = 0$
    **end if**
**end for**
**if** bidirectional edges exist **then** ▷ Check for the presence of bidirectional edges.
    *node_list* ← *nodes*($G$) ▷ Initialize node list and combinations.
    *allarcs* ← all possible combinations of any two nodes in the *node_list*
    $WL$ ← directed arcs($G$) as dataframe ▷ Retrieve directed and undirected arcs from $G$.
    *complete_arc* ← combine WL and undirected arcs($G$)
    $BL$ ← edges in *allarcs* not in *complete_arc* ▷ Identify blacklisted edges.
    *result_DAG* ← *minBIC*(*Data*, *whitelist* = $WL$, *blacklist* = $BL$), update $EAM$
**end if**
$CBN_{global} = EAM$ ▷ Set the final ensembled network.
**return** $CBN_{global}$

### S2. Performance under different network densities

This section reports additional results for simulation Category 3 under alternative variable-number and sample-size settings.

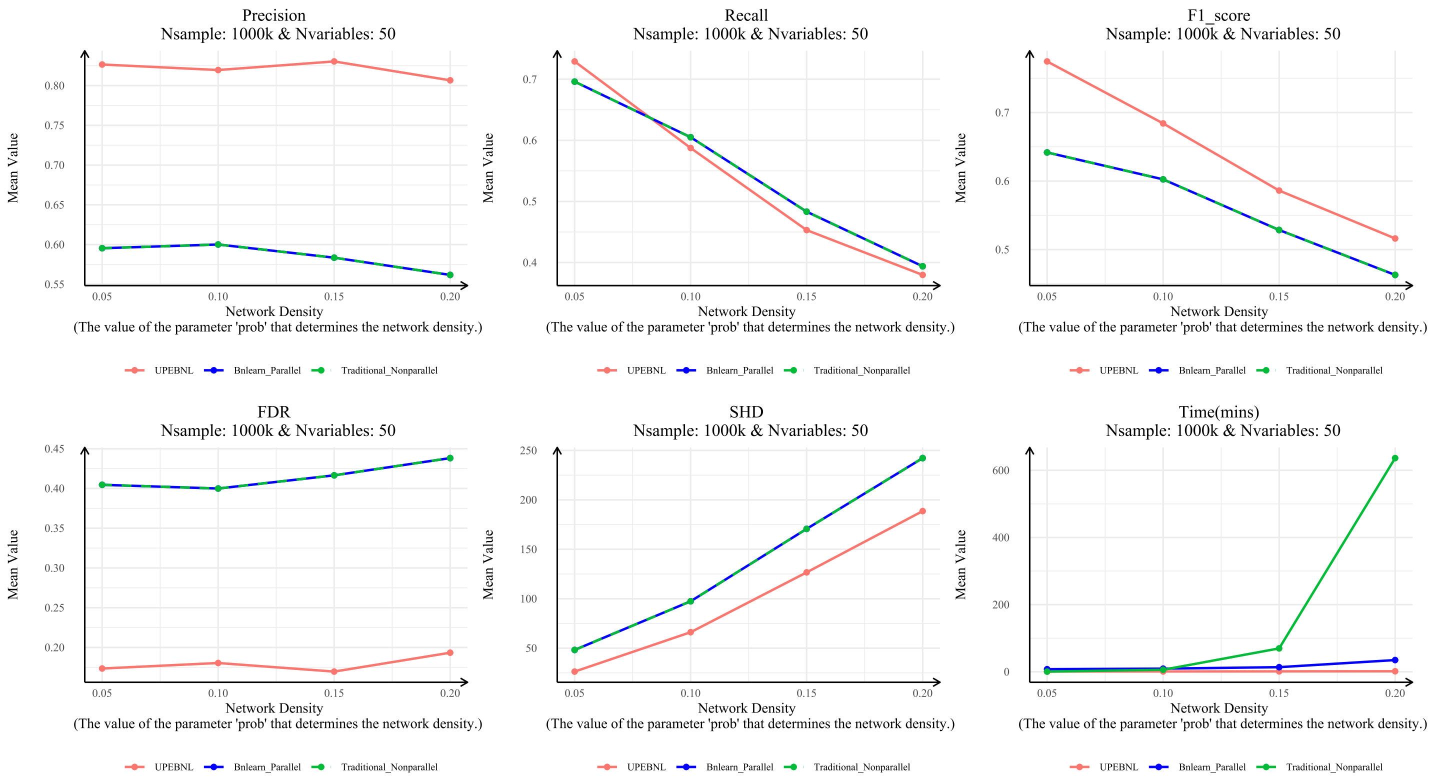


Figure S1: **Structured causal knowledge discovery under varying causal-dependency complexity with 1 million samples and 50 variables.**

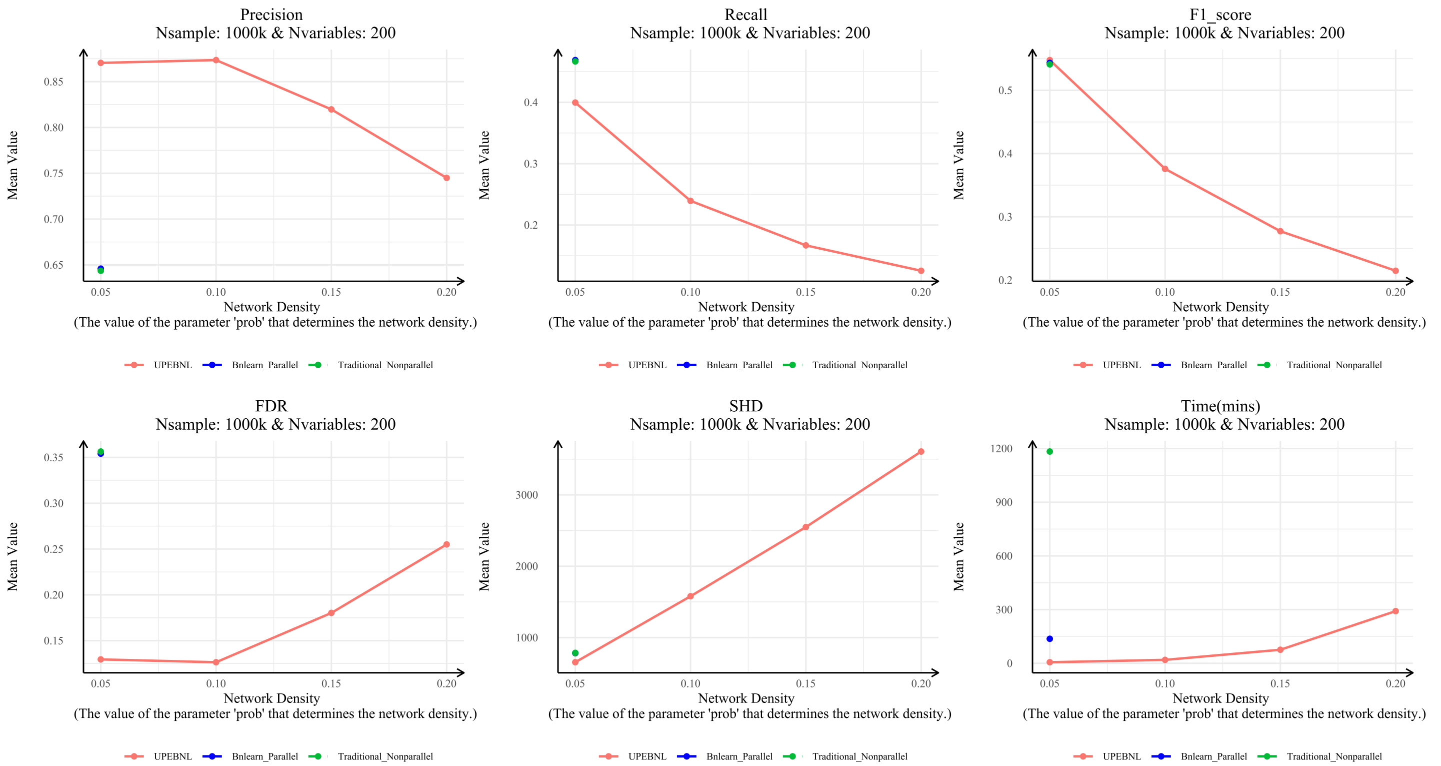


Figure S2: **Structured causal knowledge discovery under varying causal-dependency complexity with 1 million samples and 200 variables.**

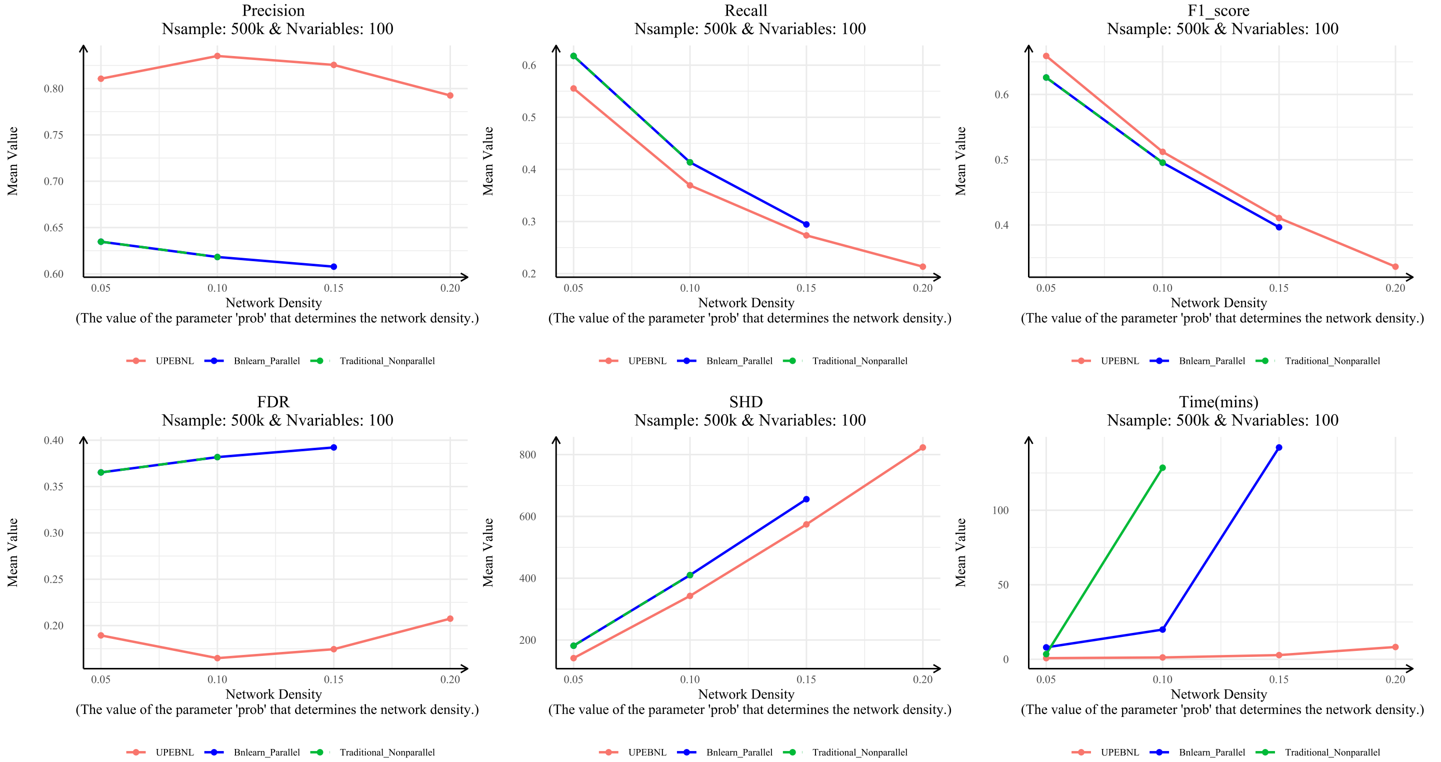


Figure S3: **Structured causal knowledge discovery under varying causal-dependency complexity with 500,000 samples and 100 variables.**

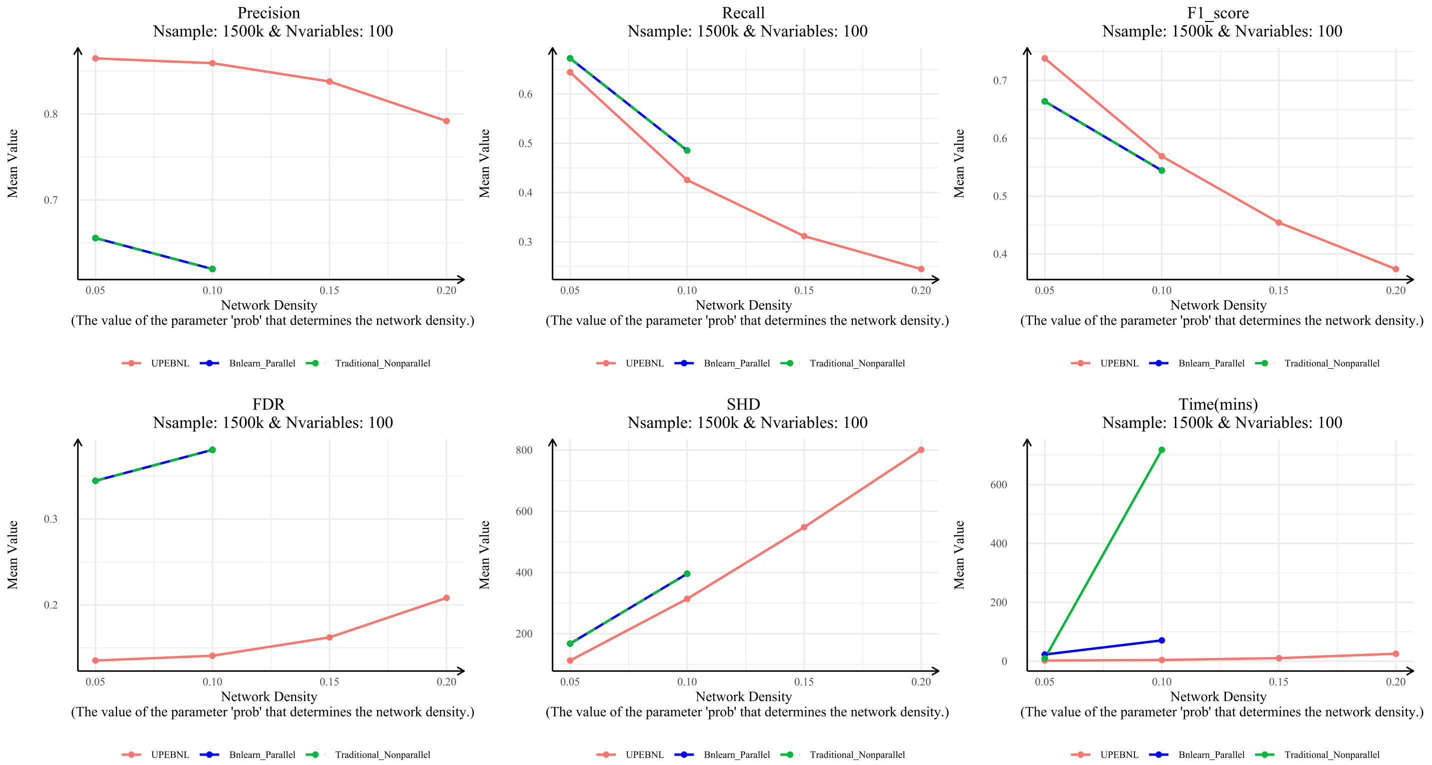


Figure S4: **Structured causal knowledge discovery under varying causal-dependency complexity with 1.5 million samples and 100 variables.**
